# Heterogeneity in pre-vaccination population immunity can contribute to variability in vaccine effectiveness estimates

**DOI:** 10.64898/2026.08.29.26361716

**Authors:** Alexander N. Pillai, Sang Woo Park, Marc Lipsitch, Benjamin J. Cowling, Sarah Cobey

## Abstract

Vaccine effectiveness (VE) estimates can vary widely between years and populations, even for the same vaccine. Estimated VE is known to be sensitive to susceptible depletion and differences in pre-vaccination infection risk between vaccinated and unvaccinated populations. However, how variation in pre-vaccination risk within and between the two groups affects VE estimates over time remains unclear. This uncertainty is especially important given negative VE estimates. We investigated the difference between estimated VE and true vaccine protection considering continuous distributions of pre-vaccination infection risk under three scenarios. When the vaccinated and unvaccinated populations differ in their mean risk, estimated VE can be higher or lower than true vaccine protection. Similar patterns arise when both populations share identical means but different risk distributions. Finally, if infection-derived immunity lasts longer than vaccine protection, annual VE estimates can vary by tens of percentage points between years despite constant true vaccine protection. These theoretical results underscore that VE studies estimate contrasting risk between vaccinated and unvaccinated individuals in a particular time and place, and VE estimates can vary counterintuitively between years and populations even with constant vaccine-induced protection. Explaining variability in estimated VE thus requires a more complete understanding of populations’ distributions of infection risk.

## Introduction

Estimates of vaccine effectiveness (VE) vary between years, locations, and populations, even for the same vaccine. For example, influenza VE estimates vary by tens of percentage points between populations in different geographic regions even when the vaccine strains are well-matched to circulating strains^1,2^. Similarly, estimates of malaria vaccine efficacy have varied across locations with different malaria parasite prevalences in the same clinical trial^3,4^. When evaluating a vaccine, the causal quantity we are often interested in is its individual-level protection against infection or disease, known as vaccine direct effects^5–10^. Estimating vaccine direct effects is challenging, particularly when randomized trials are infeasible. Instead, we commonly estimate VE using observational studies^11^, adjusting for confounders such as age, calendar time, and underlying health conditions^12,13^.

The observational design underlying VE estimates complicates their interpretation. Halloran et al.^8^ note that a common implicit assumption is that VE attempts to estimate vaccine protection as it would be observed under a hypothetical randomized controlled trial, i.e., vaccine efficacy. Under this assumption, vaccine effectiveness and efficacy both attempt to estimate vaccine direct effects^5–10^. More correctly, VE studies estimate the contrast of the risks in vaccinated and unvaccinated individuals in a particular real-world population in which individuals do not randomly choose to vaccinate^5,8,14^. VE estimates therefore reflect properties of the population as well as the vaccine, and may be biased estimates of the causal quantity of interest, i.e., vaccine direct effects (henceforth true vaccine protection). Consequently, it is important to understand how population-specific characteristics contribute to bias in VE estimates.

Without randomization, populations can differ in infection risk before vaccination. An individual’s pre-vaccination infection risk is determined by immunity, behavior, and the environment and is often ignored in simple theoretical models of estimated VE^11,15,16^. Models that have explicitly incorporated pre-vaccination risk have assumed that its distribution in vaccinated and unvaccinated populations is identical^17–21^ or that the two groups differ only in their mean pre-vaccination risk^16,22–24^. Occasionally the exact distributions are more varied but obscured^24–26^.

These models have shown that even when vaccinated and unvaccinated populations’ pre-vaccination risk distributions are identical, individual variability in infection risk can cause VE estimates to vary during an epidemic. Highly susceptible individuals become infected and immune earlier in an epidemic and will be more common in the unvaccinated population, assuming a protective vaccine^15,16,18,27–30^. As a result, the unvaccinated incidence rate decays quickly due to the depletion of susceptible unvaccinated individuals^15^. In contrast, the vaccinated incidence rate starts lower than in the unvaccinated population due to vaccine protection. If vaccines confer imperfect (“leaky”) protection, the vaccinated incidence rate also slows more gradually compared to the unvaccinated incidence rate^15^. Consequently, the difference over time in the two populations’ incidence rates, also called differential depletion of susceptibles^16,28,30^, can cause estimated VE to decrease even when true vaccine protection is constant^15,16,18,20,21,27,29,31^. If vaccine protection actually wanes over time, the effects of susceptible depletion on VE estimates become less predictable^19,32^. Models investigating the impact of differential depletion of susceptibles on estimated VE have only considered the impact within effectively a single epidemic, raising questions about the expected long-term dynamics of susceptible depletion and estimated VE.

Current models suggest the potential for substantial variation in the direction and magnitude of VE estimates with changes in population risk. Prior models have shown that differences in pre-vaccination risk can cause estimated VE to be less than the vaccine’s true protection and even negative, implying increased infection risk in the vaccinated population relative to the unvaccinated population^22,23,26^, but differential risk can sometimes instead cause VE estimates to exceed true vaccine protection^22,24,25^. Factors affecting infection risk, such as population immunity and behavior, can change over time. However, it remains unclear how the risk distributions interact with differential depletion of susceptibles and contribute to variability in VE estimates.

We systematically investigate how different distributions of pre-vaccination infection risk in vaccinated and unvaccinated populations can affect VE estimates within and across epidemics. We incorporate continuous pre-vaccination risk distributions for both populations into analytical VE models and evaluate estimated VE’s sensitivity to the distributions’ means and heterogeneity. Our synthesis of new and prior work shows how divergent immune dynamics in vaccinated and unvaccinated populations can cause annual VE estimates, such as those published for influenza vaccines, to vary despite constant true vaccine protection. Lastly, we discuss how improved measures of infection risk would help us understand variation in estimated VE across time and space and estimate true vaccine protection.

## Methods

Pre-vaccination susceptibility *ϵ* is defined as an individual’s risk of infection before vaccination and the start of an epidemic and multiplicatively scales the infection hazard *λ*. For example, an individual with pre-vaccination susceptibility *ϵ* = 2 is twice as susceptible as an individual with pre-vaccination susceptibility *ϵ* = 1, for the same infection hazard. Pre-vaccination susceptibility in the vaccinated and unvaccinated populations (*ϵ*_v_ and *ϵ*_u_ respectively) are continuous random variables following gamma distributions. Each distribution is described by its mean 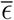 and its shape parameter *α*, and we assumed that the two distributions can have different means and shape parameters, reflecting possible differences in pre-vaccination susceptibility between vaccinated and unvaccinated populations. The shape parameter controls the level of heterogeneity and the distribution’s coefficient of variation 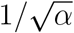. Smaller shape parameters correspond to higher heterogeneity in pre-vaccination susceptibility between individuals.

We modeled the direct effects of vaccination against infection in a partially vaccinated population, solving for the fraction of the vaccinated and unvaccinated populations that are susceptible to infection over time (see Supplemental Information S.1.2 for full derivations). For simplicity, we assumed that the infection hazard *λ* was exogenous and time-invariant, meaning that we did not model transmission between individuals. Prior work has shown that differential depletion of susceptibles affects estimates of VE similarly even with a time-varying infection hazard^16^. Vaccination generates leaky protection, and the vaccine’s direct effects, *θ*_0_, also multiplicatively scale the exogenous infection hazard. Pre-vaccination susceptibility and vaccine direct effects independently affect the infection hazard. All vaccination occurs before the epidemic begins, and vaccine direct effects are constant over time (see Supplemental Information S.1.2.4 and S.2.2 for models with waning vaccine protection). True vaccine protection was calculated as (1 *− θ*_0_) *×* 100%.

We derived analytical VE estimates from the ratios of instantaneous incidence rates 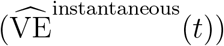, cumulative attack rates 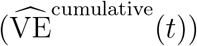, or average instantaneous incidence rates 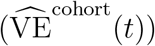 between the vaccinated and unvaccinated populations. VE is estimated as one minus the infection risk or rate ratio between the vaccinated and unvaccinated populations. For 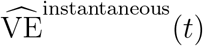, the risk ratio compares the probabilities that a vaccinated and unvaccinated individual is infected at a given time (Equation 1a). For 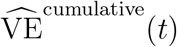, we derived the risk ratio using the probability that an individual was infected before a given time (Equation 1b). Lastly, 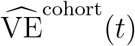 estimates VE under a cohort study design (Equation 1c). Average instantaneous incidence rates equal the cumulative attack rates divided by the expected numbers of person-time at risk in the two populations^15^, with expected person-time at risk calculated by integrating the susceptible fraction over time. VE estimates are expressed in terms of the exogenous infection hazard *λ*, vaccine direct effects *θ*_0_, and the pre-vaccination susceptibility distributions’ means (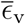 and 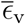) and shape parameters (*α*_v_ and *α*_u_),

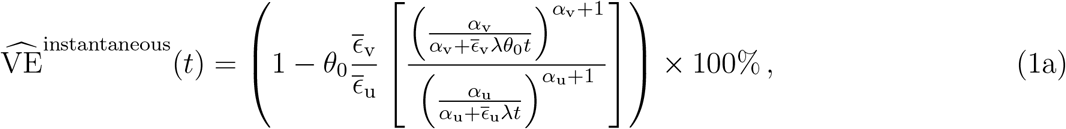

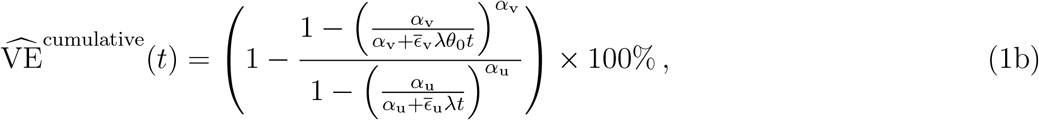

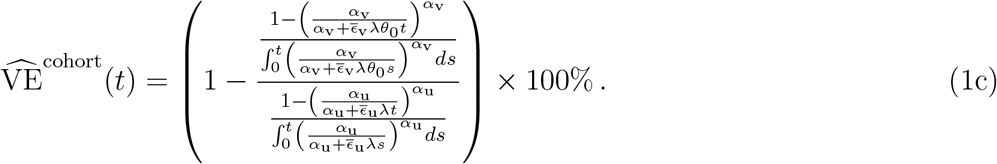

To compare our results with estimates of VE from commonly used test-negative design studies^13,33^, we also estimated VE using unconditional and conditional logistic regression, 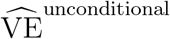 and 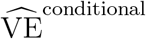 respectively, adjusting or matching for time in two-week intervals^13,33^. Unless otherwise stated, 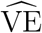 refers to our estimate using cumulative attack rates 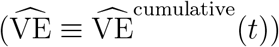 (Equation 1b).

In the first two sections of the Results, we evaluate how changing pre-vaccination susceptibility distributions affect the difference between 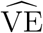 and true vaccine protection within a single epidemic. We first analyze differences in means between the vaccinated and unvaccinated distributions, holding the shape parameters constant. Then we vary the shape parameters while holding the means constant.

In the final section, we show results from an individual-based simulation (Supplemental Information S.1.4) to evaluate how 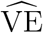 can change across epidemics. After initialization, the susceptibility distributions arise endogenously from the boosting and waning of protection after infections and vaccinations, changing from year to year. All software for this work is available at https://github.com/cobeylab/heterogeneity_in_pre-vax_immunity_contributes_to_ve_variability.

## Results

### VE estimates are sensitive to the mean pre-vaccination susceptibilities of vaccinated and unvaccinated populations

Differences in mean pre-vaccination susceptibility between the vaccinated and unvaccinated populations cause VE estimated toward the end of the epidemic, or final 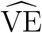, to differ from true vaccine protection^16,22–24^. Previous work has shown that differences in pre-vaccination susceptibility lead to different rates of susceptible depletion in each population. Generally, differential depletion of susceptibles causes VE estimates to decline over time^15,16,18,21^. To evaluate whether pre-vaccination susceptibility impacts VE estimates early in an epidemic, before appreciable susceptible depletion occurs, we investigated 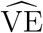 estimates at the limit of time approaching zero, defined as starting 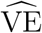:

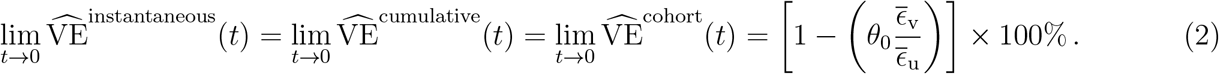

Equation 2 follows from Equations 1a–1c (Supplemental Information S.2.1). Note that when vaccinated and unvaccinated populations have identical mean pre-vaccination susceptibilities 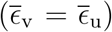, starting 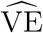 equals true vaccine protection ([1 *− θ*_0_] *×* 100%) regardless of the mean. Furthermore, heterogeneity in pre-vaccination susceptibility (*α*_v_ and *α*_u_) does not affect starting 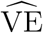.

When the vaccinated and unvaccinated populations have different mean pre-vaccination susceptibilities 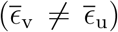, starting 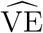 deviates from the vaccine’s true protection (Equation 2). The effect’s direction depends on the ratio of mean pre-vaccination susceptibilities between the two populations, 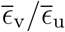. When the vaccinated population has lower mean pre-vaccination susceptibility than the unvaccinated population, starting 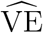 exceeds true vaccine protection (blue tiles in Figure 1 A). In the opposite case, starting 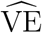 is lower than the vaccine’s true protection (orange tiles in Figure 1 A). When the unvaccinated population has much lower mean pre-vaccination susceptibility than the vaccinated population 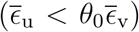, starting 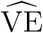 can be negative (e.g., top-left highlighted tile in Figure 1 A).

**Figure 1.**
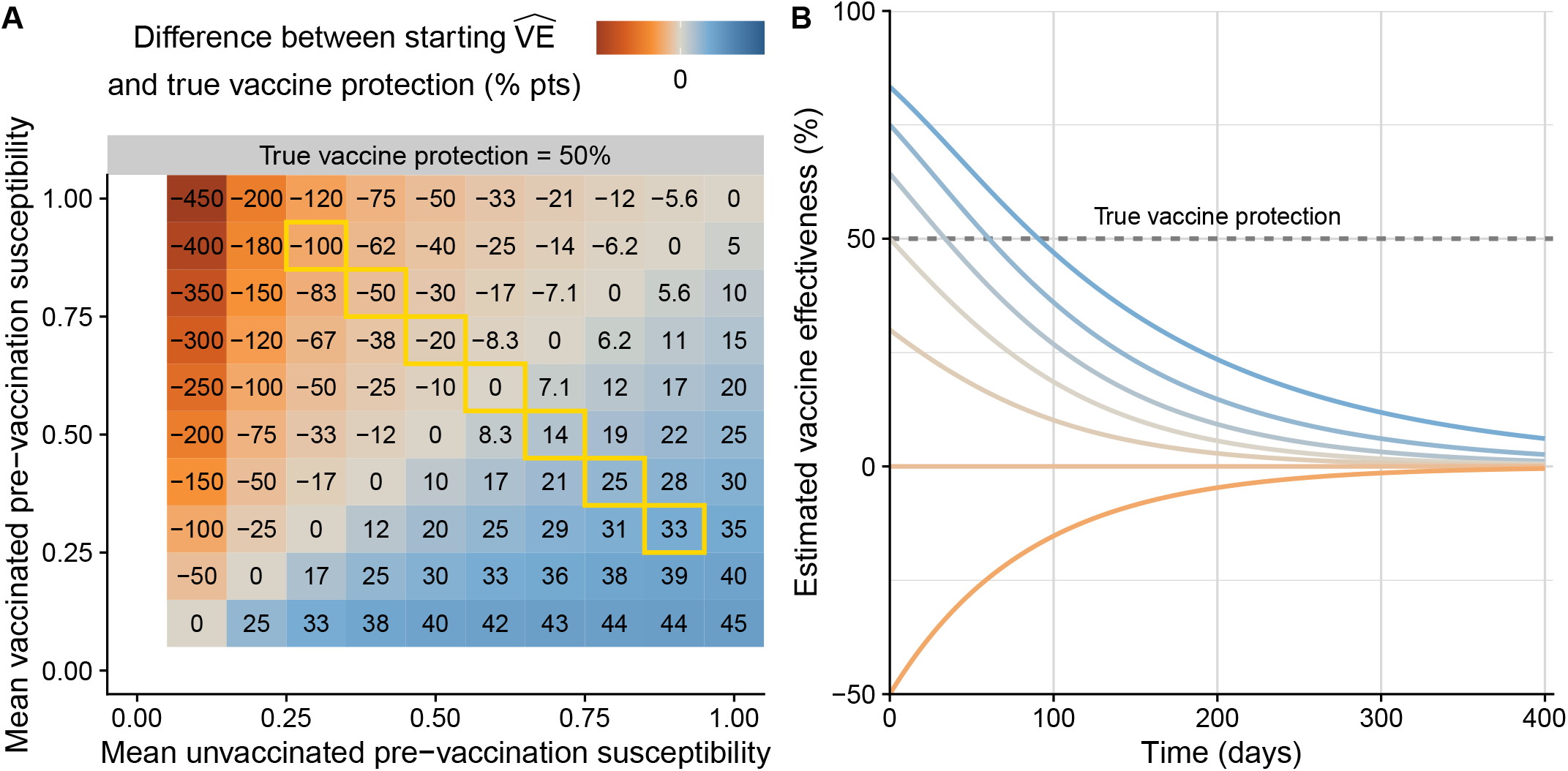
Starting 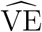 differs from true vaccine protection when vaccinated and unvaccinated populations have different mean pre-vaccination susceptibilities. (A) The vaccinated and unvaccinated populations have continuously distributed pre-vaccination susceptibility with different means but identical shape parameters corresponding to a highly homogeneous distribution (*α*_v_ = *α*_u_ = 20). Each cell shows the difference between starting 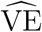 and true vaccine protection (50% in this case). Darker blue cells show when starting 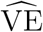 exceeds true vaccine protection and darker orange corresponds to starting 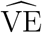 that is lower than true protection. (B) Example 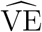 trajectories colored by the difference between starting 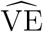 and true vaccine protection. The curves correspond to the yellow highlighted cells in A, and the *t* = 0 intercepts of each curve equals the starting 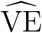 for each highlighted cell. For these example trajectories, the exogenous infection hazard *λ* = 0.05.

Trajectories of 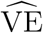 converge toward zero and start higher or lower depending on the mean pre-vaccination susceptibility ratio (Figure 1 B). When pre-vaccination susceptibility is very homogeneous and true vaccine protection is constant, 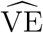 decreases towards zero when starting 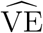 is positive^15,16^ and increases towards zero when starting 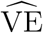 is negative. If, however, vaccine protection wanes, different pre-vaccination means in the vaccinated and unvaccinated populations can result in 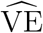 crossing zero during an epidemic (Supplemental Figure S.4). Regression-based final 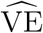 estimates are almost identical to final 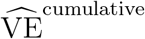 estimates, differing by less than a percentage point on average (95% interval: −1 to 1 percentage-point difference for 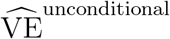 and 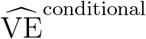). Final 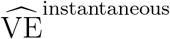 estimates also differ from 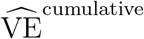 estimates by less than a percentage point on average but vary more (95% interval: −19 to 67 percentage-point difference). Higher variability in 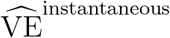 is expected since VE estimated from instantaneous incidence rates is more sensitive to differential depletion of susceptibles^16^. Final 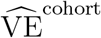 estimates are 3 percentage points lower than 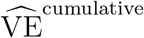 estimates on average and also show more variability (95% interval: −72 to 11 percentage-point difference). Higher variability in 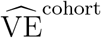 occurs because 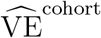 estimates are less sensitive to susceptible depletion when pre-vaccination susceptibility is less heterogeneous^15^ and remain closer to starting 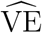.

### Heterogeneity in pre-vaccination susceptibility can cause estimated VE to exceed true vaccine protection or become negative

Heterogeneity in vaccinated and unvaccinated pre-vaccination susceptibility can also cause VE estimates to differ from true vaccine protection^18,21,27^. To investigate its effects systematically, we varied the susceptibility distributions’ shape parameters while keeping their means identical. Because the populations have identical mean pre-vaccination susceptibilities, starting 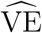 is equal to true vaccine protection. Therefore, the difference between final 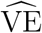 estimates and true vaccine protection reflects how different levels of heterogeneity in pre-vaccination susceptibility impacts the two populations’ infection rates over time.

When heterogeneity in pre-vaccination susceptibility is identical in the vaccinated and unvaccinated populations (*α*_v_ = *α*_u_), final 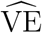 increasingly underestimates true vaccine protection as heterogeneity increases^17,18,21^. High heterogeneity means that there are more individuals at the extremes of susceptibility (Figure 2 A). Because large fractions of both populations have very low susceptibility, incidence rates in the two populations quickly become more similar, and cumulative attack rates become more parallel over time (Figure 2 B). As a result, 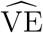 is lower across all values of true vaccine protection as heterogeneity increases (Figure 2 C).

**Figure 2.**
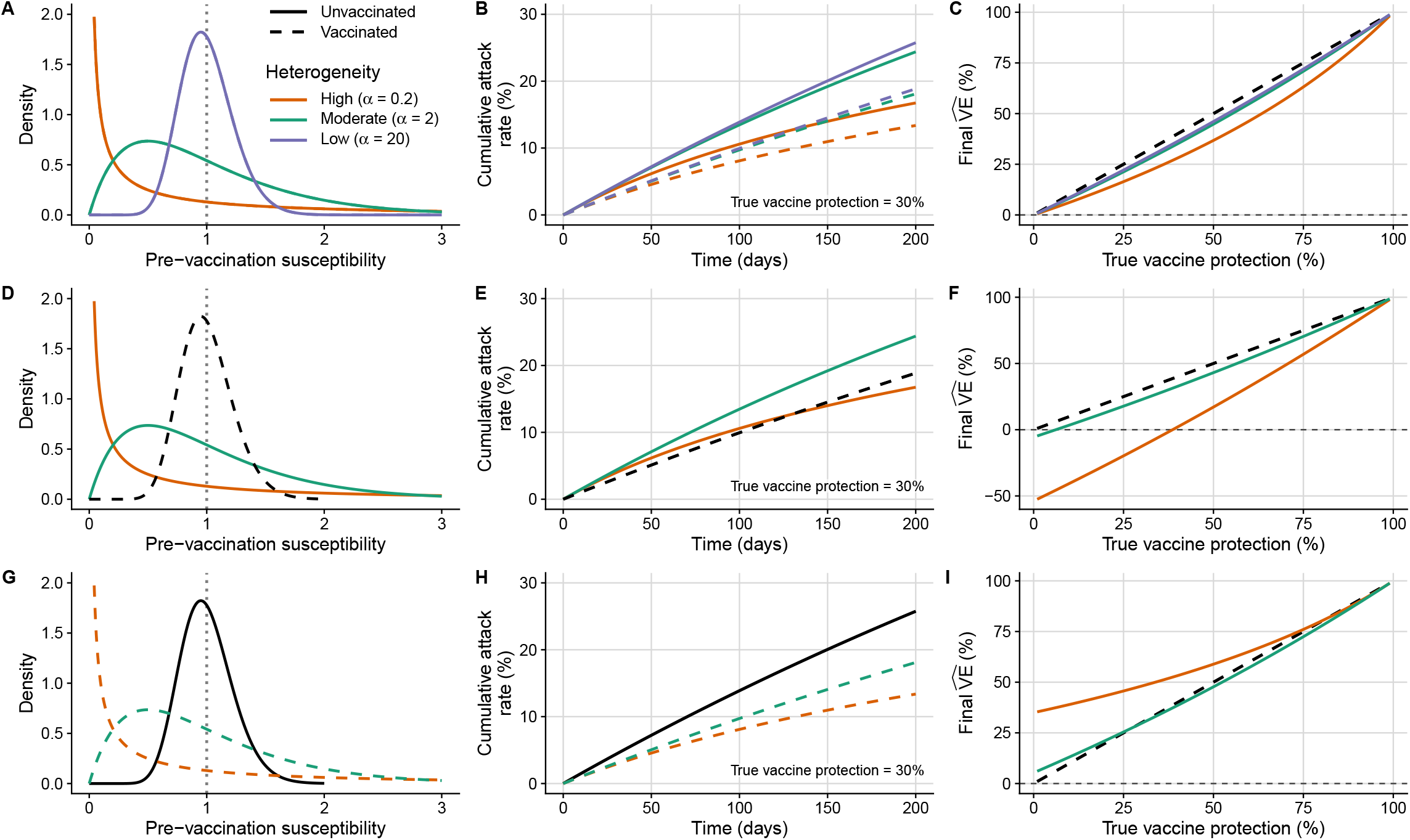
Heterogeneity in pre-vaccination susceptibility changes the rates of susceptible depletion and their effects on 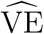. Across all scenarios, the pre-vaccination susceptibility distributions have identical means 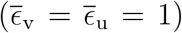, shown as vertical dotted lines in A, D, and G. The shapes of the pre-vaccination susceptibility distributions vary from low heterogeneity (*α* = 20) to high heterogeneity (*α* = 0.2). (A) Vaccinated and unvaccinated pre-vaccination susceptibility distributions have identical heterogeneity (*α*_v_ = *α*_u_). (B) Cumulative attack rates in the vaccinated and unvaccinated populations (dashed and solid curves respectively) when true vaccine protection is 30%. (C) The relationship between true vaccine protection and final 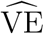. The black dashed diagonal represents the one-to-one relationship. Panels D–F show the same curves when heterogeneity is higher in the unvaccinated than the vaccinated population (*α*_v_ = 20). Black dashed curves in D and E show the vaccinated population’s pre-vaccination susceptibility distribution and cumulative attack rate. Panels G–I show curves when heterogeneity is higher in the vaccinated than in the unvaccinated population (*α*_u_ = 20). Black solid curves in G and H show the unvaccinated pre-vaccination susceptibility distribution and cumulative attack rate. For all scenarios, the exogenous infection hazard *λ* = 0.0015, and epidemics lasted for 200 days.

When heterogeneity in pre-vaccination susceptibility differs between vaccinated and unvaccinated populations (*α*_v_ ≠ *α*_u_), there is more variability in infection rates between the two populations. If pre-vaccination susceptibility is more heterogeneous in the unvaccinated than the vaccinated population (Figure 2 D), high-risk unvaccinated individuals are infected early and become immune. As the epidemic progresses, the unvaccinated incidence rate slows quickly, since the remaining unvaccinated people have low susceptibility (colored curves in Figure 2 E). In contrast, the vaccinated incidence rate changes gradually over time (black dashed curve in Figure 2 E), and final 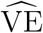 is lower than true vaccine protection and even negative (Figure 2 F). Similar dynamics apply in the opposite case: if the vaccinated population has more heterogeneous pre-vaccination susceptibility (Figure 2 G), the vaccinated incidence rate slows more quickly than in the unvaccinated population (Figure 2 H), and final 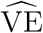 can exceed true vaccine protection (Figure 2 I). Differential heterogeneity in pre-vaccination susceptibility affects final 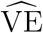 in the same direction regardless of how VE is estimated (Supplemental Figure S.8). Across all scenarios, regression-based 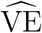 estimates are lower than 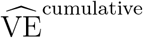 estimates by less than a percentage point on average (95% interval: −2 to 0.8 percentage-point difference for 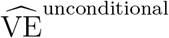 and −2 to 0.7 for 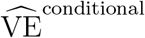), whereas 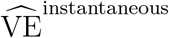 estimates are 10 percentage points lower than 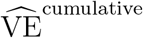 on average (95% interval: −58 to 16 percentage-point difference). Final 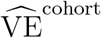 estimates are higher than 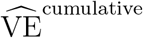 estimates by 2 percentage points on average (95% interval: −3 to 4 percentage-point difference).

### Estimated VE can change across years due to dynamics in population immunity

Combining the results of the preceding two sections, we illustrate how population immunity can drive variability in estimates of VE across annual epidemics. Using an individual-based simulation, we calculated annual 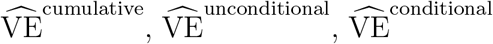 and 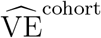 estimates in a partially vaccinated population (Supplemental Information S.1.4). We focused on the VE estimates that use infection data from the entire epidemic, since final 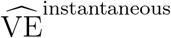 estimates are more sensitive to differential depletion of susceptibles^16^ and are uncommon in observational studies. We considered a scenario where infection-derived immune protection lasts longer than vaccine protection, assuming that vaccinated people in one year have an 85% probability of re-vaccinating in the next year, reflecting the high observed autocorrelation in annual influenza vaccination status^34,35^; vaccine-derived immune protection is constant over an epidemic and immediately lost if an individual does not vaccinate at the start of the next year^36,37^; and infection-derived immune protection wanes exponentially with a four-year half life^38^. The exogenous infection hazard was seasonally forced to produce epidemics with an approximately 25% cumulative attack rate on average, with a larger-than-normal epidemic in the fourth year (Figure 3 A). At the start of the simulation, the vaccinated and unvaccinated pre-vaccination susceptibility distributions were initialized with identical means, but the unvaccinated population started with a more heterogeneous pre-vaccination susceptibility than the vaccinated population, as could occur after a large epidemic in which more unvaccinated than vaccinated people were infected and developed protective immunity.

**Figure 3.**
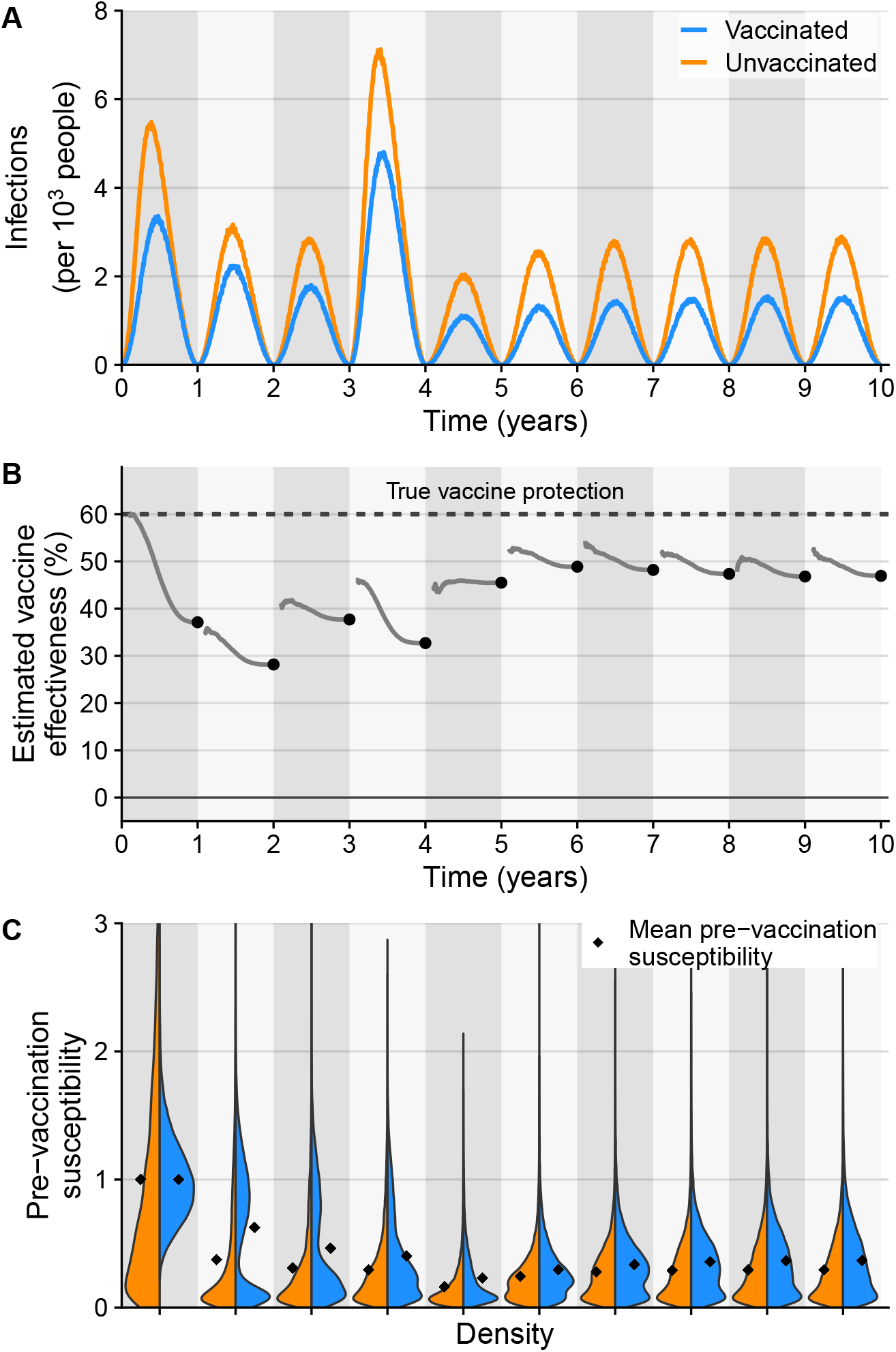
Dynamic population immunity causes year-to-year variation in 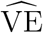 despite constant true vaccine protection. (A) Infection incidence in the vaccinated and unvaccinated populations (blue and orange respectively) per 1,000 people. Note that the infection hazard is seasonally forced, unlike in the preceding sections. (B) Time-varying 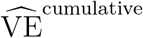 is shown in gray, and final 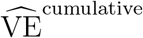 indicated by the black dot. (C) Distributions of pre-vaccination susceptibility in the vaccinated and unvaccinated populations (blue and orange respectively) at the start of each year. Black diamonds show the means of pre-vaccination susceptibility distributions.

The dynamics of population immunity cause 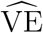 to vary by tens of percentage points over time despite no change in underlying vaccine protection. With a 60% protective vaccine, 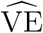 ranges approximately 10–30 percentage points below the true level of protection (Figure 3 B). In the first year, when pre-vaccination susceptibility is more heterogeneous in the unvaccinated population (Figure 3 C), 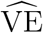 changes by over 20 percentage points during the epidemic. This occurs because a substantial fraction of the unvaccinated population has high prior immune protection, or near-zero pre-vaccination susceptibility. As a result, the cumulative attack rates in the two populations become more similar over the epidemic, similar to Figures 2 E and 2 F. After the first epidemic, mean pre-vaccination susceptibility is lower in the unvaccinated than the vaccinated population, reflecting accumulated immunity from infections. 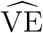 in the second year thus starts and remains 20–30 percentage points lower than true vaccine protection. In later years, there are smaller differences between starting 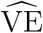 and true vaccine protection as vaccinated and unvaccinated mean pre-vaccination susceptibilities become more similar, and final 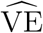 estimates stabilize near 45–50%. 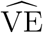 trajectories within epidemics become flatter because the pre-vaccination susceptibility distributions’ shapes are more similar between the two populations. Final annual 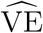 estimates are similar across different estimation methods (Supplemental Figure S.11). Regression-based final 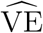 estimates are lower 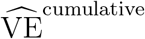 by less than one percentage point on average (95% interval: −3 to 2 percentage-point difference for 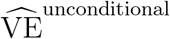 and 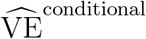). Final 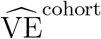 estimates are higher than 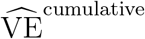 estimates by 5 percentage points on average (95% interval: 3 to 12 percentage-point difference).

## Discussion

Differences in immunity, behavior, and other infection risk factors between vaccinated and unvaccinated populations contribute to variability in VE estimates across time and space^16,22–24,26,39^, but theoretical models have yet to systematically investigate how differences in the distributions of pre-vaccination infection risk impact estimated VE within and across epidemics. We demonstrate how such differences affect susceptible depletion and the difference between 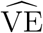 and true vaccine protection. When mean risks differ between vaccinated and unvaccinated populations, 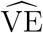 can be higher or lower than true vaccine protection throughout an epidemic. Alternatively, when the means are identical, differences in heterogeneity influence the infection rates in vaccinated and unvaccinated populations over time, causing changes to final 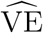 in either direction. Simulations of vaccination and infection with an influenza-like pathogen demonstrate that the dynamics of population immunity can cause annual 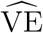 estimates to vary by tens of percentage points.

Prior theoretical work has suggested that when mean pre-vaccination risk does not vary by vaccination status, estimated VE is less than true vaccine protection due to differential depletion of susceptibles^15,16,18,20,21,27,29,31^. But we found that 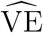 can, in fact, exceed true vaccine protection if the vaccinated population has sufficiently heterogeneous pre-vaccination risk compared to the unvaccinated population. The risk distributions in different groups could compound or counteract any effects of differences in mean pre-vaccination risk on 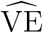. The net effect of differential pre-vaccination risk on final 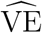 estimates is therefore challenging to predict without information on these risk distributions.

Differences in pre-vaccination risk distributions affect 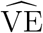 regardless of how it is estimated. VE estimated with instantaneous incidence rates, 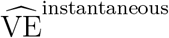, changes in similar directions as 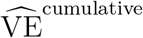 but typically diverges more from true vaccine protection. VE estimated from instantaneous incidence rates is more sensitive to differential depletion of susceptibles, resulting in larger differences between 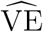 and true vaccine protection at the end of an epidemic^16^. Variation in final 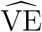 is similar in direction and magnitude regardless of whether VE is estimated with logistic regression or cumulative attack rates. VE estimated under a cohort study design also varies in similar directions as 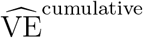 estimates, but the magnitude of variation is sometimes larger because 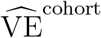is less sensitive to differential depletion of susceptibles^15^ and remains closer to starting 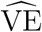. It is therefore likely that variability in pre-vaccination infection risk across individuals and time affects real-world observational VE estimates, since the distributions’ effects are not limited to any one study design.

Our analysis carries several limitations. We assumed that pre-vaccination infection risk and vaccine-derived protection impact the infection hazard independently. However, prior immunity affects vaccine responses and protection^36,39–42^, which could lead to different risk distributions than investigated here. We assumed that vaccination protects all vaccinees as the same multiple of their individual infection hazard, but more biologically precise models are possible. The simulations assumed no protection from prior-year vaccination, but residual protection may exist^36,37^. All models assume that vaccination occurs before the epidemic starts, which results in two limitations. First, if vaccination overlaps the start of the epidemic, highly susceptible people may be infected before vaccination, altering the rate of susceptible depletion^19^. Second, if vaccination coverage varies during an epidemic, regression analyses that adjust for calendar time could deviate further from our theoretical VE estimates based on cumulative attack rates^12,43–46^. Simulated epidemics approach an equilibrium because only immune waning and vaccination status affect susceptibility in the model, and the average exogenous infection hazard is mostly constant. Antigenic evolution, changes in behavior, and changes in true vaccine protection and coverage can make real-world epidemic dynamics less stable. Relatedly, explicitly modeling transmission may be more appropriate when considering the indirect effects of widespread vaccination^9^.

An implication of our results is that when the distributions of pre-vaccination infection risk are unknown, comparisons of VE estimates in different time periods and populations require careful interpretation. In Figure 3, we illustrate how population immune dynamics generate year-to-year variation in 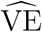. Without knowing the time-varying risk distributions, we may falsely conclude that true vaccine protection ([1 *− θ*_0_] *×* 100%) changes each year. Differential distributions of pre-vaccination risk may similarly contribute to reduced estimates of VE in people vaccinated several years in a row relative to infrequent vaccinees^40,47–49^ or in some age or population groups^50,51^. As mentioned previously, there may also be real changes to vaccine direct effects between years, which could interact in complex ways with risk distributions. For example, antigenic evolution might create a mismatch between vaccine and epidemic strains that affects some people more than others, depending on which viral epitopes their antibodies target^50,51^.

Comparisons of VE estimates could therefore be strengthened by better measures of factors other than vaccination that contribute to infection risk in vaccinated and unvaccinated populations, which could help distinguish vaccine direct effects from other differences in susceptibility. Correlates of infection risk— including infection and vaccination history^35–38,40,52,53^, immune measures^54–58^, sociodemographic characteristics^59–61^, environmental conditions^62,63^, and time-varying individual behaviors^64–66^—remain poorly quantified even for well studied pathogens such as influenza and SARS-CoV-2. Immune mediators of protection could additionally improve estimates of rates of vaccine waning. Overall, improved measures of infection risk could enhance assessments of vaccination’s effect in different populations and clarify why estimates of VE vary across time and space.

## Acknowledgments

We thank Graham Northrup, Lauren McGough, Stefano Allesina, Greg Dwyer, and Isabel Rodriguez-Barraquer for their comments. Natural language processing tools driven by artificial intelligence were not used to derive models, write code, or process data, but they were used to perform reviews of the code and manuscript after final editing.

## Conflicts of interest

B.J.C. has consulted for AstraZeneca, GlaxoSmithKline, Moderna, Novavax, Pfizer, Roche, Sanofi Pasteur and Seqirus. M.L. has participated in a one-day unpaid consultation with Merck on a vaccine unrelated to respiratory infections and performed compensated consulting work for the Gates Foundation and Analysis Group on unrelated matters. All other authors report no potential conflicts of interest.

## Funding

This study was supported by the National Institute of Allergy and Infectious Diseases (NIAID) Collaborative Influenza Vaccine Innovation Centers (CIVICs) (contract 75N93019C00051). The content of this article is solely the responsibility of the authors and does not necessarily represent the official views of the National Institutes of Health (NIH). A.P. was supported by the Quantitative Ecology GAANN training grant (grant P200A210054) from the Department of Education. S.W.P. was supported by the New Faculty Startup Fund from Seoul National University.

## Data availability

Code for all analytical and simulation models is available at https://github.com/cobeylab/heterogeneity_in_pre-vax_immunity_contributes_to_ve_variability. Analytical and regression analyses were performed in R version 4.3.3. The simulation model was implemented in Julia version 1.10.2.

## References

[1] Edward A Belongia et al. “Variable influenza vaccine effectiveness by subtype: a systematic review and meta-analysis of test-negative design studies”. In: The Lancet Infectious Diseases 16.8 (Aug. 1, 2016), pp. 942–951. doi: 10.1016/S1473-3099(16)00129-8.

[2] G. N. Okoli et al. “Variable seasonal influenza vaccine effectiveness across geographical regions, age groups and levels of vaccine antigenic similarity with circulating virus strains: A systematic review and meta-analysis of the evidence from test-negative design studies after the 2009/10 influenza pandemic”. In: Vaccine 39.8 (Feb. 22, 2021), pp. 1225–1240. doi: 10.1016/j.vaccine.2021.01.032.

[3] Philip Bejon et al. “Efficacy of RTS,S malaria vaccines: individual-participant pooled analysis of phase 2 data”. In: The Lancet Infectious Diseases 13.4 (Apr. 1, 2013), pp. 319–327. doi: 10.1016/S1473-3099(13)70005-7.

[4] Ally Olotu et al. “Seven-Year Efficacy of RTS,S/AS01 Malaria Vaccine among Young African Children”. In: New England Journal of Medicine 374.26 (June 30, 2016). eprint: https://www.nejm.org/doi/p pp. 2519–2529. doi: 10.1056/NEJMoa1515257.

[5] Sander Greenland and Ralph R Frerichs. “On Measures and Models for the Effectiveness of Vaccines and Vaccination Programmes”. In: International Journal of Epidemiology 17.2 (June 1, 1988), pp. 456–463. doi: 10.1093/ije/17.2.456.

[6] Claudio J Struchiner et al. “The Behaviour of Common Measures of Association Used to Assess a Vaccination Programme under Complex Disease Transmission Patterns—A Computer Simulation Study of Malaria Vaccines”. In: International Journal of Epidemiology 19.1 (Mar. 1, 1990), pp. 187–196. doi: 10.1093/ije/19.1.187.

[7] Michael Haber, Ira M Longini Jr, and M Elizabeth Halloran. “Measures of the Effects of Vaccination in a Randomly Mixing Population”. In: International Journal of Epidemiology 20.1 (Mar. 1, 1991), pp. 300–310. doi: 10.1093/ije/20.1.300.

[8] M. Elizabeth Halloran et al. “Direct and Indirect Effects in Vaccine Efficacy and Effectiveness”. In: American Journal of Epidemiology 133.4 (Feb. 15, 1991), pp. 323–331. doi: 10.1093/oxfordjournals.aje.a115884.

[9] M. E. Halloran, C. J. Struchiner, and I. M. Longini. “Study Designs for Evaluating Different Efficacy and Effectiveness Aspects of Vaccines”. In: American Journal of Epidemiology 146.10 (Nov. 15, 1997), pp. 789–803. doi: 10.1093/oxfordjournals.aje.a009196.

[10] M. E. Halloran, I. M. Longini, and C. J. Struchiner. “Design and interpretation of vaccine field studies”. In: Epidemiologic Reviews 21.1 (1999), pp. 73–88. doi: 10.1093/oxfordjournals.epirev.a017990.

[11] Michael L. Jackson and Jennifer C. Nelson. “The test-negative design for estimating influenza vaccine effectiveness”. In: Vaccine 31.17 (Apr. 19, 2013), pp. 2165–2168. doi: 10.1016/j.vaccine.2013.02.053.

[12] Sheena G. Sullivan, Eric J. Tchetgen Tchetgen, and Benjamin J. Cowling. “Theoretical Basis of the Test-Negative Study Design for Assessment of Influenza Vaccine Effectiveness”. In: American Journal of Epidemiology 184.5 (Sept. 1, 2016), pp. 345–353. doi: 10.1093/aje/kww064.

[13] Huiying Chua et al. “The Use of Test-negative Controls to Monitor Vaccine Effectiveness: A Systematic Review of Methodology”. In: Epidemiology 31.1 (Jan. 2020), p. 43. doi: 10.1097/EDE.0000000000001116.

[14] George W. Comstock. “Vaccine evaluation by case-control or prospective studies”. In: American Journal of Epidemiology 131.2 (Feb. 1, 1990), pp. 205–207. doi: 10.1093/oxfordjournals.aje.a115490.

[15] P G Smith, L C Rodrigues, and P E M Fine. “Assessment of the Protective Efficacy of Vaccines against Common Diseases Using Case-Control and Cohort Studies”. In: International Journal of Epidemiology 13.1 (1984), pp. 87–93. doi: 10.1093/ije/13.1.87.

[16] Joseph A Lewnard et al. “Measurement of Vaccine Direct Effects Under the Test-Negative Design”. In: American Journal of Epidemiology 187.12 (Dec. 1, 2018), pp. 2686–2697. doi: 10.1093/aje/kwy163.

[17] M. Elizabeth Halloran, Ira M. Longini Jr., and Claudio J. Struchiner. “Estimability and Interpretation of Vaccine Efficacy Using Frailty Mixing Models”. In: American Journal of Epidemiology 144.1 (July 1, 1996), pp. 83–97. doi: 10.1093/oxfordjournals.aje.a008858.

[18] Michael T. White et al. “Heterogeneity in malaria exposure and vaccine response: implications for the interpretation of vaccine efficacy trials”. In: Malaria Journal 9.1 (Mar. 23, 2010), p. 82. doi: 10.1186/1475-2875-9-82.

[19] M. Lipsitch et al. “Depletion-of-susceptibles bias in influenza vaccine waning studies: how to ensure robust results”. In: Epidemiology & Infection 147 (Jan. 2019), e306. doi: 10.1017/S0950268819001961.

[20] Rebecca Kahn et al. “Identifying and Alleviating Bias Due to Differential Depletion of Susceptible People in Postmarketing Evaluations of COVID-19 Vaccines”. In: American Journal of Epidemiology 191.5 (Mar. 24, 2022), pp. 800–811. doi: 10.1093/aje/kwac015.

[21] Ariel Nikas, Hasan Ahmed, and Veronika I. Zarnitsyna. “Competing Heterogeneities in Vaccine Effectiveness Estimation”. In: Vaccines 11.8 (Aug. 2023). Number: 8, p. 1312. doi: 10.3390/vaccines11081312.

[22] Joseph A. Lewnard et al. “Theoretical Framework for Retrospective Studies of the Effectiveness of SARS-CoV-2 Vaccines”. In: Epidemiology (Cambridge, Mass.) 32.4 (July 2021), pp. 508–517. doi: 10.1097/EDE.0000000000001366.

[23] Korryn Bodner et al. “Testing Whether Higher Contact Among the Vaccinated Can Be a Mechanism for Observed Negative Vaccine Effectiveness”. In: American Journal of Epidemiology 192.8 (Aug. 4, 2023), pp. 1335–1340. doi: 10.1093/aje/kwad055.

[24] Kylie E. C. Ainslie et al. “On the bias of estimates of influenza vaccine effectiveness from test–negative studies”. In: Vaccine 35.52 (Dec. 19, 2017), pp. 7297–7301. doi: 10.1016/j.vaccine.2017.10.107.

[25] Ivo M. Foppa et al. “Vaccination history as a confounder of studies of influenza vaccine effectiveness”. In: Vaccine: X 1 (Apr. 11, 2019), p. 100008. doi: 10.1016/j.jvacx.2019.100008.

[26] Ryan E. Wiegand et al. “Bias and negative values of COVID-19 vaccine effectiveness estimates from a test-negative design without controlling for prior SARS-CoV-2 infection”. In: Nature Communications 15 (Nov. 20, 2024), p. 10062. doi: 10.1038/s41467-024-54404-w.

[27] Marc Lipsitch. “Challenges of Vaccine Effectiveness and Waning Studies”. In: Clinical Infectious Diseases: An Official Publication of the Infectious Diseases Society of America 68.10 (May 15, 2019), pp. 1631–1633. doi: 10.1093/cid/ciy773.

[28] Claudio J. Struchiner et al. “Malaria vaccines: lessons from field trials”. In: Cadernos de Saúde Pública 10 (1994), S310–S326. doi: 10.1590/S0102-311X1994000800009.

[29] Edward Goldstein et al. “Temporally varying relative risks for infectious diseases: implications for infectious disease control”. In: Epidemiology (Cambridge, Mass.) 28.1 (Jan. 2017), pp. 136–144. doi: 10.1097/EDE.0000000000000571.

[30] Miguel A. Hernán. “The Hazards of Hazard Ratios”. In: Epidemiology (Cambridge, Mass.) 21.1 (Jan. 2010), pp. 13–15. doi: 10.1097/EDE.0b013e3181c1ea43.

[31] Eunha Shim and Alison P. Galvani. “Distinguishing vaccine efficacy and effectiveness”. In: Vaccine 30.47 (Oct. 19, 2012), pp. 6700–6705. doi: 10.1016/j.vaccine.2012.08.045.

[32] Rebecca Kahn et al. “Examining Bias From Differential Depletion of Susceptibles in Vaccine Effectiveness Estimates in Settings of Waning”. In: American Journal of Epidemiology 193.1 (Sept. 28, 2023), pp. 232–234. doi: 10.1093/aje/kwad191.

[33] Sheena G Sullivan, Shuo Feng, and Benjamin J Cowling. “Potential of the test-negative design for measuring influenza vaccine effectiveness: a systematic review”. In: Expert Review of Vaccines 13.12 (Dec. 1, 2014). eprint: https://doi.org/10.1586/14760584.2014.966695, pp. 1571–1591. doi: 10.1586/14760584.2014.966695.

[34] May P. S. Yeung, Frank L.Y. Lam, and Richard Coker. “Factors associated with the uptake of seasonal influenza vaccination in adults: a systematic review”. In: Journal of Public Health 38.4 (Dec. 2, 2016), pp. 746–753. doi: 10.1093/pubmed/fdv194.

[35] Philip Arevalo et al. “Earliest infections predict the age distribution of seasonal influenza A cases”. In: eLife 9 (July 7, 2020). Ed. by Ben S Cooper et al., e50060. doi: 10.7554/eLife.50060.

[36] Edward A. Belongia et al. “Repeated annual influenza vaccination and vaccine effectiveness: review of evidence”. In: Expert Review of Vaccines 16.7 (July 3, 2017), pp. 723–736. doi: 10.1080/14760584.2017.1334554.

[37] Elenor Jones-Gray et al. “Does repeated influenza vaccination attenuate effectiveness? A systematic review and meta-analysis”. In: The Lancet Respiratory Medicine 11.1 (Jan. 1, 2023), pp. 27–44. doi: 10.1016/S2213-2600(22)00266-1.

[38] Sylvia Ranjeva et al. “Age-specific differences in the dynamics of protective immunity to influenza”. In: Nature Communications 10.1 (Apr. 10, 2019), p. 1660. doi: 10.1038/s41467-019-09652-6.

[39] Joseph A. Lewnard and Sarah Cobey. “Immune History and Influenza Vaccine Effectiveness”. In: Vaccines 6.2 (June 2018). Number: 2, p. 28. doi: 10.3390/vaccines6020028.

[40] Qifang Bi et al. “Reduced Effectiveness of Repeat Influenza Vaccination: Distinguishing Among Within-Season Waning, Recent Clinical Infection, and Subclinical Infection”. In: The Journal of Infectious Diseases 230.6 (Dec. 15, 2024), pp. 1309–1318. doi: 10.1093/infdis/jiae220.

[41] Niklas Bobrovitz et al. “Protective effectiveness of previous SARS-CoV-2 infection and hybrid immunity against the omicron variant and severe disease: a systematic review and meta-regression”. In: The Lancet Infectious Diseases 23.5 (May 1, 2023), pp. 556–567. doi: 10.1016/S1473-3099(22)00801-5.

[42] Tim K Tsang et al. “Prior infections and effectiveness of SARS-CoV-2 vaccine in test-negative studies: a systematic review and meta-analysis”. In: American Journal of Epidemiology 193.12 (Dec. 2, 2024), pp. 1868–1881. doi: 10.1093/aje/kwae142.

[43] Peter Jacoby and Heath Kelly. “Is it necessary to adjust for calendar time in a test negative design?: Responding to: Jackson ML, Nelson JC. The test negative design for estimating influenza vaccine effectiveness. Vaccine 2013;31(April (17)):2165–8”. In: Vaccine 32.25 (May 23, 2014), p. 2942. doi: 10.1016/j.vaccine.2013.08.048.

[44] H. S. Bond, S. G. Sullivan, and B. J. Cowling. “Regression approaches in the test-negative study design for assessment of influenza vaccine effectiveness”. In: Epidemiology & Infection 144.8 (June 2016), pp. 1601–1611. doi: 10.1017/S095026881500309X.

[45] Jozef J. P. Nauta. “Adjusting for calendar time in a TND influenza study”. In: Epidemiology & Infection 144.12 (Sept. 2016), pp. 2689–2690. doi: 10.1017/S0950268816000832.

[46] Natalie E Dean, M Elizabeth Halloran, and Ira M Longini Jr. “Temporal Confounding in the Test-Negative Design”. In: American Journal of Epidemiology 189.11 (Nov. 2, 2020), pp. 1402–1407. doi: 10.1093/aje/kwaa084.

[47] T. W. Hoskins et al. “Assessment of Inactivated Influenza-A Vaccine After Three Outbreaks of Influenza a at Christ’s Hospital”. In: The Lancet. Originally published as Volume 1, Issue 8106 313.8106 (Jan. 6, 1979), pp. 33–35. doi: 10.1016/S0140-6736(79)90468-9.

[48] Danuta M. Skowronski et al. “Association between the 2008–09 Seasonal Influenza Vaccine and Pandemic H1N1 Illness during Spring–Summer 2009: Four Observational Studies from Canada”. In: PLOS Medicine 7.4 (Apr. 6, 2010), e1000258. doi: 10.1371/journal.pmed.1000258.

[49] Benjamin J. Cowling et al. “Reply to Skowronski”. In: Clinical Infectious Diseases 52.6 (Mar. 15, 2011), pp. 832–833. doi: 10.1093/cid/cir041.

[50] Danuta M. Skowronski et al. “Paradoxical clade- and age-specific vaccine effectiveness during the 2018/19 influenza A(H3N2) epidemic in Canada: potential imprint-regulated effect of vaccine (I-REV)”. In: Eurosurveillance 24.46 (Nov. 14, 2019), p. 1900585. doi: 10.2807/1560-7917.ES.2019.24.46.1900585.

[51] Brendan Flannery et al. “Spread of Antigenically Drifted Influenza A(H3N2) Viruses and Vaccine Effectiveness in the United States During the 2018–2019 Season”. In: The Journal of Infectious Diseases 221.1 (Jan. 1, 2020), pp. 8–15. doi: 10.1093/infdis/jiz543.

[52] Katelyn M. Gostic et al. “Potent protection against H5N1 and H7N9 influenza via childhood hemagglutinin imprinting”. In: Science (New York, N.Y.) 354.6313 (Nov. 11, 2016), pp. 722–726. doi: 10.1126/science.aag1322.

[53] Nobuo Saito et al. “Negative impact of prior influenza vaccination on current influenza vaccination among people infected and not infected in prior season: A test-negative case-control study in Japan”. In: Vaccine 35.4 (Jan. 23, 2017), pp. 687–693. doi: 10.1016/j.vaccine.2016.11.024.

[54] John S. Tsang et al. “Global analyses of human immune variation reveal baseline predictors of post-vaccination responses”. In: Cell 157.2 (Apr. 10, 2014), pp. 499–513. doi: 10.1016/j.cell.2014.03.031.

[55] Leah C. Katzelnick et al. “Antibody-dependent enhancement of severe dengue disease in humans”. In: Science 358.6365 (Nov. 17, 2017), pp. 929–932. doi: 10.1126/science.aan6836.

[56] Kangchon Kim et al. “Measures of Population Immunity Can Predict the Dominant Clade of Influenza A (H3N2) in the 2017–2018 Season and Reveal Age-Associated Differences in Susceptibility and Antibody-Binding Specificity”. In: Influenza and Other Respiratory Viruses 18.11 (2024). eprint: https://onlinelibrary.wiley.com/doi/pdf/10.1111/irv.70033, e70033. doi: 10.1111/irv.70033.

[57] Caroline Kikawa et al. “High-throughput neutralization measurements correlate strongly with evolutionary success of human influenza strains”. In: eLife 14 (Feb. 23, 2026). Ed. by Bryan D Bryson and Aleksandra M Walczak, RP106811. doi: 10.7554/eLife.106811.

[58] Weijia Xiong et al. “Measuring population immunity against influenza using individual antibody titres: a multicountry, retrospective observational study”. In: The Lancet Infectious Diseases (Apr. 17, 2026). doi: 10.1016/S1473-3099(26)00061-7.

[59] Q. Sue Huang et al. “Risk Factors and Attack Rates of Seasonal Influenza Infection: Results of the Southern Hemisphere Influenza and Vaccine Effectiveness Research and Surveillance (SHIVERS) Seroepidemiologic Cohort Study”. In: The Journal of infectious diseases 219.3 (Jan. 9, 2019), pp. 347–357. doi: 10.1093/infdis/jiy443.

[60] Casey M. Zipfel, Vittoria Colizza, and Shweta Bansal. “Health inequities in influenza transmission and surveillance”. In: PLOS Computational Biology 17.3 (Mar. 11, 2021), e1008642. doi: 10.1371/journal.pcbi.1008642.

[61] Jacob A. Udell et al. “Clinical risk, sociodemographic factors, and SARS-CoV-2 infection over time in Ontario, Canada”. In: Scientific Reports 12.1 (June 24, 2022), p. 10534. doi: 10.1038/s41598-022-13598-z.

[62] Juliana C. Taube et al. “Characterising non-household contact patterns relevant to respiratory transmission in the USA: analysis of a cross-sectional survey”. In: The Lancet Digital Health 7.8 (Aug. 1, 2025). doi: 10.1016/j.landig.2025.100888.

[63] Nicolas Banholzer et al. “The relative contribution of close-proximity contacts, shared classroom exposure and indoor air quality to respiratory virus transmission in schools”. In: Nature Communications 16.1 (Nov. 27, 2025), p. 11678. doi: 10.1038/s41467-025-66719-3.

[64] Shweta Bansal, Bryan T Grenfell, and Lauren Ancel Meyers. “When individual behaviour matters: homogeneous and network models in epidemiology”. In: Journal of The Royal Society Interface 4.16 (Oct. 22, 2007), pp. 879–891. doi: 10.1098/rsif.2007.1100.

[65] Gonzalo M. Vazquez-Prokopec et al. “Using GPS Technology to Quantify Human Mobility, Dynamic Contacts and Infectious Disease Dynamics in a Resource-Poor Urban Environment”. In: PLOS ONE 8.4 (Apr. 8, 2013), e58802. doi: 10.1371/journal.pone.0058802.

[66] Adam J. Kucharski et al. “The Contribution of Social Behaviour to the Transmission of Influenza A in a Human Population”. In: PLOS Pathogens 10.6 (June 26, 2014), e1004206. doi: 10.1371/journal.ppat.1004206.

