## Supplemental Information for "Heterogeneity in pre-vaccination population immunity can contribute to variability in vaccine effectiveness estimates"

**S.3 Supplemental figures** **19**

### S.1 Full description of methods

#### S.1.1 Test-negative design exposure odds ratio under simplifying assumptions

Here we show how the exposure odds ratio from test-negative design studies (TND OR) can simplify into the infection risk ratio conditional on vaccination status, following prior theoretical models[1,2]. We then use the infection risk ratio as the basis for our $\widehat{\mathrm{VE}}$ models in the next section.

The TND OR requires the expected counts of vaccinated and unvaccinated test-positive and test-negative infections $X_{vi}$, where the subscripts $v$ and $i$ indicate vaccination and infection status, respectively (e.g., $X_{11}$ represents the number of vaccinated test-positive infections). We define the model's random variables in Supplemental Table S.1, and we let $N$ be the total population size.

| Random variable | Description | Values |
|---|---|---|
| $O$ | Observation status | $o \in \{0 = \text{unobserved infection}, 1 = \text{observed infection}\}$ |
| $H$ | Healthcare-seeking behavior | $h \in \{0 = \text{does not seek care}, 1 = \text{seeks care}\}$ |
| $S$ | Symptom status | $s \in \{0 = \text{asymptomatic}, 1 = \text{symptomatic}\}$ |
| $I$ | Infection status | $i \in \{0 = \text{infected by test-negative pathogen}, 1 = \text{infected by test-positive pathogen}\}$ |
| $V$ | Vaccination status | $v \in \{0 = \text{unvaccinated}, 1 = \text{vaccinated}\}$ |

**Table S.1:** Random variables in TND OR test-positive and test-negative infection count expressions.

The expected number of infections depends on the joint probability of observation, infection, and vaccination status, $\Pr(O = o, I = i, V = v)$. We decomposed the joint probability using the conditional probabilities of detection and infection. The expected number of vaccinated test-positive infections, $X_{11}$, becomes

$$\begin{aligned} X_{11} &= \Pr(O = 1, I = 1, V = 1)N \\ &= \Pr(O = 1|I = 1, V = 1)\Pr(I = 1|V = 1)\Pr(V = 1)N\,. \end{aligned} \tag{S.1}$$

We similarly simplified the remaining expected infection counts:

$$X_{01} = \Pr(O = 1|I = 1, V = 0)\Pr(I = 1|V = 0)\Pr(V = 0)N\,, \tag{S.2}$$
$$X_{10} = \Pr(O = 1|I = 0, V = 1)\Pr(I = 0|V = 1)\Pr(V = 1)N\,, \tag{S.3}$$
$$X_{00} = \Pr(O = 1|I = 0, V = 0)\Pr(I = 0|V = 0)\Pr(V = 0)N\,. \tag{S.4}$$

We followed standard TND assumptions to simplify the expected infection count expressions. We assumed that vaccination has no effect on the probability of infection from test-negative pathogens, $\Pr(I = 0|V = v) = \Pr(I = 0)$. We also assumed that the probability of an infection being observed combines the probability of an infection developing symptoms ($S$) and the probability that an individual with a symptomatic infection seeks medical care ($H$). We therefore rewrote the first terms in Supplemental Equations S.1–S.4 as $\Pr(O = 1|I = i, V = v) = \Pr(H = 1, S = 1|I = i, V = v)$. We made the same assumptions as prior theoretical TND models[1]:

1. The probabilities of seeking care and developing symptoms are independent, $\Pr(H = 1, S = 1|I = i, V = v) = \Pr(H = 1|I = i, V = v)\Pr(S = 1|I = i, V = v)$.

2. The probability of seeking care is independent of infection etiology, $\Pr(H = 1|I = i, V = v) = \Pr(H = 1|V = v)$.

3. The probability of developing symptoms is independent of vaccination status, $\Pr(S = 1|I = i, V = v) = \Pr(S = 1|I = i)$.

We therefore simplified the expected case and control counts in terms of $H$, $S$, $I$, and $V$:

$$X_{11} = \Pr(H = 1|V = 1) \Pr(S = 1|I = 1) \Pr(I = 1|V = 1) \Pr(V = 1)N\,, \tag{S.5}$$
$$X_{01} = \Pr(H = 1|V = 0) \Pr(S = 1|I = 1) \Pr(I = 1|V = 0) \Pr(V = 0)N\,, \tag{S.6}$$
$$X_{10} = \Pr(H = 1|V = 1) \Pr(S = 1|I = 0) \Pr(I = 0) \Pr(V = 1)N\,, \tag{S.7}$$
$$X_{00} = \Pr(H = 1|V = 0) \Pr(S = 1|I = 0) \Pr(I = 0) \Pr(V = 0)N\,. \tag{S.8}$$

The odds of vaccination in the test-positive and test-negative infections are $X_{11}/X_{01}$ and $X_{10}/X_{00}$, respectively:

$$\begin{aligned}\frac{X_{11}}{X_{01}} &= \frac{\Pr(H = 1|V = 1) \Pr(S = 1|I = 1) \Pr(I = 1|V = 1) \Pr(V = 1)N}{\Pr(H = 1|V = 0) \Pr(S = 1|I = 1) \Pr(I = 1|V = 0) \Pr(V = 0)N} \\ &= \frac{\Pr(H = 1|V = 1) \Pr(I = 1|V = 1) \Pr(V = 1)}{\Pr(H = 1|V = 0) \Pr(I = 1|V = 0) \Pr(V = 0)}\,,\end{aligned} \tag{S.9}$$

$$\begin{aligned}\frac{X_{10}}{X_{00}} &= \frac{\Pr(H = 1|V = 1) \Pr(S = 1|I = 0) \Pr(I = 0) \Pr(V = 1)N}{\Pr(H = 1|V = 0) \Pr(S = 1|I = 0) \Pr(I = 0) \Pr(V = 0)N} \\ &= \frac{\Pr(H = 1|V = 1) \Pr(V = 1)}{\Pr(H = 1|V = 0) \Pr(V = 0)}\,.\end{aligned} \tag{S.10}$$

The TND OR is the ratio of $X_{11}/X_{01}$ and $X_{10}/X_{00}$ and simplifies to the infection risk ratio between the vaccinated and unvaccinated populations,

$$\begin{aligned}\mathrm{OR}_{\mathrm{TND}} = \frac{\frac{X_{11}}{X_{01}}}{\frac{X_{10}}{X_{00}}} &= \frac{X_{11}\,X_{00}}{X_{01}\,X_{10}} \\ &= \frac{[\Pr(H = 1|V = 1) \Pr(I = 1|V = 1) \Pr(V = 1)]\,[\Pr(H = 1|V = 0) \Pr(V = 0)]}{[\Pr(H = 1|V = 0) \Pr(I = 1|V = 0) \Pr(V = 0)]\,[\Pr(H = 1|V = 1) \Pr(V = 1)]} \\ &= \frac{\Pr(I = 1|V = 1)}{\Pr(I = 1|V = 0)}\,.\end{aligned} \tag{S.11}$$

##### S.1.2 Derivation of VE estimate models

In the main-text estimates, we assumed that vaccine direct effects are constant, such that $\theta(t) = \theta_0$, $\forall t$. To evaluate the impact of waning vaccine protection, we assumed that vaccine direct effects decayed exponentially from a starting protection, $1 - \theta_0$, to zero with a half life $\eta$. We therefore modeled vaccine direct effects as

$$\theta(t) = \begin{cases} \theta_0\,, & \text{without waning,} \\ 1 - \left[(1-\theta_0)\,e^{-t\left(\frac{\ln 2}{\eta}\right)}\right]\,, & \text{with waning.}\end{cases} \tag{S.12}$$

We also defined $\Theta(t) = \int_0^t \theta(s)\,ds$ to be the total vaccine effect over the time interval $[0, t]$ such that

$$\Theta(t) = \begin{cases} \theta_0 t\,, & \text{without waning,} \\ t + \frac{(2^{-t/\eta})(2^{t/\eta}-1)(\eta)(\theta_0-1)}{\ln(2)}\,, & \text{with waning.}\end{cases} \tag{S.13}$$

The expression for total vaccine effects with waning was solved using Mathematica.

As described in the main text, the population distribution of pre-vaccination susceptibility follows a gamma distribution. Subscripts v and u correspond to the vaccinated ($V = 1$) and unvaccinated ($V = 0$) populations. Vaccinated and unvaccinated pre-vaccination susceptibilities ($\epsilon_\mathrm{v}$ and $\epsilon_\mathrm{u}$ respectively) follow distinct distributions described by a shape parameter, $\alpha$, and mean, $\bar{\epsilon} = \alpha/\beta$, where $\beta$ is the distribution's rate parameter. We assumed that the exogenous infection hazard is constant over time ($\lambda(t) = \lambda$, $\forall t$) and is independently scaled by pre-vaccination susceptibility and vaccine direct effects.

We modeled how susceptibility changes in the vaccinated and unvaccinated populations over time, for a given pre-vaccination susceptibility value:

$$\frac{\mathrm{d}S_\mathrm{u}}{\mathrm{d}t}(t, \epsilon_\mathrm{u}) = -\epsilon_\mathrm{u}\lambda S_\mathrm{u}(t, \epsilon_\mathrm{u}) \qquad \epsilon_\mathrm{u} \sim \mathrm{Gamma}(\alpha_\mathrm{u},\, \beta_\mathrm{u})\,, \tag{S.14}$$

$$\frac{\mathrm{d}S_\mathrm{v}}{\mathrm{d}t}(t, \epsilon_\mathrm{v}) = -\theta(t)\epsilon_\mathrm{v}\lambda S_\mathrm{v}(t, \epsilon_\mathrm{v}) \qquad \epsilon_\mathrm{v} \sim \mathrm{Gamma}(\alpha_\mathrm{v},\, \beta_\mathrm{v})\,. \tag{S.15}$$

We then solved for the proportion of each population susceptible at a given time, assuming all individuals start susceptible to infection ($S_\mathrm{u}(0,\, \epsilon_\mathrm{u}) = S_\mathrm{v}(0,\, \epsilon_\mathrm{v}) = 1$):

$$S_\mathrm{u}(t, \epsilon_\mathrm{u}) = S_\mathrm{u}(0, \epsilon_\mathrm{u})e^{-\epsilon_\mathrm{u}\lambda t} = e^{-\epsilon_\mathrm{u}\lambda t}\,, \tag{S.16}$$

$$S_\mathrm{v}(t, \epsilon_\mathrm{v}) = S_\mathrm{v}(0, \epsilon_\mathrm{v})e^{\int_0^t -\theta(s)\epsilon_\mathrm{v}\lambda\, ds} = e^{-\epsilon_\mathrm{v}\lambda\Theta(t)}\,. \tag{S.17}$$

Next, we integrated over the pre-vaccination susceptibility distribution, $f_\mathrm{Gamma}(\epsilon)$. We present a generic example of this derivation. Let $\tilde{\lambda} = \lambda t$ such that $e^{-\epsilon\lambda t} = e^{-\epsilon\tilde{\lambda}}$, simplifying the exponential terms. To start, we substituted the gamma distribution's probability density function into the integrand and simplified:

$$\begin{aligned} S(t) &= \int_0^\infty e^{-\epsilon\tilde{\lambda}} f_\mathrm{Gamma}(\epsilon;\, \alpha, \beta)\, d\epsilon \\ &= \int_0^\infty e^{-\epsilon\tilde{\lambda}} \left[\frac{\beta^\alpha e^{-\beta\epsilon}\epsilon^{\alpha-1}}{\Gamma(\alpha)}\right] d\epsilon \\ &= \frac{\beta^\alpha}{\Gamma(\alpha)} \int_0^\infty e^{-\epsilon(\beta+\tilde{\lambda})}\epsilon^{\alpha-1}\, d\epsilon\,. \end{aligned} \tag{S.18}$$

We multiplied and divided the integrand by $\frac{(\beta+\tilde{\lambda})^\alpha}{\Gamma(\alpha)}$. This becomes the normalizing constant for the integrand, transforming it into a valid gamma distribution probability density function that integrates to one:

$$\begin{aligned} S(t) &= \frac{\beta^\alpha}{\Gamma(\alpha)} \int_0^\infty \left[\frac{(\beta+\tilde{\lambda})^\alpha e^{-\epsilon(\beta+\tilde{\lambda})}\epsilon^{\alpha-1}}{\Gamma(\alpha)}\right]\left[\frac{\Gamma(\alpha)}{(\beta+\tilde{\lambda})^\alpha}\right] d\epsilon \\ &= \left[\frac{\beta^\alpha}{\Gamma(\alpha)}\right]\left[\frac{\Gamma(\alpha)}{(\beta+\tilde{\lambda})^\alpha}\right] \int_0^\infty f_\mathrm{Gamma}(\epsilon;\, \alpha, \beta+\tilde{\lambda})\, d\epsilon \\ &= \left[\frac{\beta}{(\beta+\tilde{\lambda})}\right]^\alpha. \end{aligned} \tag{S.19}$$

We further simplified the expression using the fact that the gamma distribution's rate parameter equals the ratio of the shape parameter and the mean ($\beta = \alpha\,/\,\bar{\epsilon}$):

$$S(t) = \left(\frac{\alpha}{\alpha + \bar{\epsilon}\tilde{\lambda}}\right)^\alpha. \tag{S.20}$$

For the unvaccinated population, $\tilde{\lambda} = \lambda t$, and for the vaccinated population, $\tilde{\lambda} = \lambda\Theta(t)$. With these substitutions, we solved for the susceptible fractions in both populations:

$$S_{\mathrm{u}}(t) = \int_0^\infty e^{-\epsilon_{\mathrm{u}}\lambda t} f_{\mathrm{Gamma}}(\epsilon_{\mathrm{u}}; \alpha_{\mathrm{u}}, \beta_{\mathrm{u}})\, d\epsilon_{\mathrm{u}} = \left(\frac{\alpha_{\mathrm{u}}}{\alpha_{\mathrm{u}} + \bar{\epsilon}_{\mathrm{u}}\lambda t}\right)^{\alpha_{\mathrm{u}}}, \tag{S.21}$$

$$S_{\mathrm{v}}(t) = \int_0^\infty e^{-\epsilon_{\mathrm{v}}\lambda \Theta(t)} f_{\mathrm{Gamma}}(\epsilon_{\mathrm{v}}; \alpha_{\mathrm{v}}, \beta_{\mathrm{v}})\, d\epsilon_{\mathrm{v}} = \left(\frac{\alpha_{\mathrm{v}}}{\alpha_{\mathrm{v}} + \bar{\epsilon}_{\mathrm{v}}\lambda \Theta(t)}\right)^{\alpha_{\mathrm{v}}}. \tag{S.22}$$

We also modeled the instantaneous incidence rates in the vaccinated and unvaccinated populations for a given pre-vaccination susceptibility value,

$$\frac{\mathrm{d}I_{\mathrm{u}}}{\mathrm{d}t}(t, \epsilon_{\mathrm{u}}) = \epsilon_{\mathrm{u}}\lambda S_{\mathrm{u}}(t, \epsilon_{\mathrm{u}}) = \epsilon_{\mathrm{u}}\lambda e^{-\epsilon_{\mathrm{u}}\lambda t}, \tag{S.23}$$

$$\frac{\mathrm{d}I_{\mathrm{v}}}{\mathrm{d}t}(t, \epsilon_{\mathrm{v}}) = \theta(t)\epsilon_{\mathrm{v}}\lambda S_{\mathrm{v}}(t, \epsilon_{\mathrm{v}}) = \theta(t)\epsilon_{\mathrm{v}}\lambda e^{-\epsilon_{\mathrm{v}}\lambda\Theta(t)}. \tag{S.24}$$

We again integrated over the pre-vaccination susceptibility distributions and present a generic case of the derivation. Consider a generic version of the instantaneous incidence rate, $\epsilon\lambda a e^{-\epsilon\lambda b}$. Let $\tilde{\lambda} = \lambda b$ and $\lambda' = \lambda a$, such that the expression becomes $\epsilon\lambda' e^{-\epsilon\tilde{\lambda}}$. We began by substituting the gamma distribution's probability density function into the integrand alongside the simplified instantaneous incidence rate expression:

$$\begin{aligned}
\frac{\mathrm{d}I}{\mathrm{d}t}(t) &= \int_0^\infty \epsilon\lambda' e^{-\epsilon\tilde{\lambda}} f_{\mathrm{Gamma}}(\epsilon; \alpha, \beta)\, d\epsilon \\
&= \int_0^\infty \epsilon\lambda' e^{-\epsilon\tilde{\lambda}} \left[\frac{\beta^\alpha e^{-\beta\epsilon}\epsilon^{\alpha-1}}{\Gamma(\alpha)}\right] d\epsilon \\
&= \frac{\lambda'\beta^\alpha}{\Gamma(\alpha)} \int_0^\infty e^{-\epsilon(\beta+\tilde{\lambda})}\epsilon^{(\alpha+1)-1}\, d\epsilon.
\end{aligned} \tag{S.25}$$

We multiplied and divided the integrand by $\frac{(\beta+\tilde{\lambda})^{(\alpha+1)}}{\Gamma(\alpha+1)}$, turning the integrand into a valid gamma probability density function that integrates to one:

$$\begin{aligned}
\frac{\mathrm{d}I}{\mathrm{d}t}(t) &= \frac{\lambda'\beta^\alpha}{\Gamma(\alpha)} \int_0^\infty \left[\frac{(\beta+\tilde{\lambda})^{(\alpha+1)} e^{-\epsilon(\beta+\tilde{\lambda})}\epsilon^{(\alpha+1)-1}}{\Gamma(\alpha+1)}\right] \left[\frac{\Gamma(\alpha+1)}{(\beta+\tilde{\lambda})^{(\alpha+1)}}\right] d\epsilon \\
&= \frac{\lambda'\beta^\alpha\Gamma(\alpha+1)}{(\beta+\tilde{\lambda})^{(\alpha+1)}\Gamma(\alpha)} \int_0^\infty f_{\mathrm{Gamma}}(\epsilon; \alpha+1, \beta+\tilde{\lambda})\, d\epsilon.
\end{aligned} \tag{S.26}$$

We further simplified the expression using a property of gamma functions, that $\Gamma(\alpha+1) = \alpha\Gamma(\alpha)$:

$$\begin{aligned}
\frac{\mathrm{d}I}{\mathrm{d}t}(t) &= \frac{\lambda'\beta^\alpha\alpha\Gamma(\alpha)}{(\beta+\tilde{\lambda})^{(\alpha+1)}\Gamma(\alpha)} \\
&= \frac{\lambda'\beta^\alpha\alpha}{(\beta+\tilde{\lambda})^{(\alpha+1)}} \\
&= \frac{\lambda'\alpha}{\beta+\tilde{\lambda}}\left(\frac{\beta}{\beta+\tilde{\lambda}}\right)^\alpha.
\end{aligned} \tag{S.27}$$

The right-hand term is identical to Supplemental Equation S.19 and can similarly be simplified using

$\beta = \alpha/\bar{\epsilon}$. We also used this relationship to simplify the left-hand term:

$$\begin{aligned}\frac{\mathrm{d}I}{\mathrm{d}t}(t) &= \frac{\lambda'\alpha}{\frac{\alpha}{\bar{\epsilon}} + \tilde{\lambda}}\left(\frac{\alpha}{\alpha + \bar{\epsilon}\tilde{\lambda}}\right)^{\alpha} \\ &= \frac{\lambda'\bar{\epsilon}\alpha}{\alpha + \bar{\epsilon}\tilde{\lambda}}\left(\frac{\alpha}{\alpha + \bar{\epsilon}\tilde{\lambda}}\right)^{\alpha} \\ &= \lambda'\bar{\epsilon}\left(\frac{\alpha}{\alpha + \bar{\epsilon}\tilde{\lambda}}\right)^{\alpha+1}.\end{aligned} \tag{S.28}$$

For the unvaccinated population, $\tilde{\lambda} = \lambda t$ and $\lambda' = \lambda$. For the vaccinated population, $\tilde{\lambda} = \lambda\Theta(t)$ and $\lambda' = \lambda\theta(t)$. We therefore solved for the instantaneous incidence rates in the two populations:

$$\frac{\mathrm{d}I_{\mathrm{u}}}{\mathrm{d}t}(t) = \int_0^{\infty} \epsilon_{\mathrm{u}}\lambda e^{-\epsilon_{\mathrm{u}}\lambda t} f_{\mathrm{Gamma}}(\epsilon_{\mathrm{u}}; \alpha_{\mathrm{u}}, \beta_{\mathrm{u}})\, d\epsilon_{\mathrm{u}} = \lambda\bar{\epsilon}_{\mathrm{u}}\left(\frac{\alpha_{\mathrm{u}}}{\alpha_{\mathrm{u}} + \bar{\epsilon}_{\mathrm{u}}\lambda t}\right)^{\alpha_{\mathrm{u}}+1}, \tag{S.29}$$

$$\frac{\mathrm{d}I_{\mathrm{v}}}{\mathrm{d}t}(t) = \int_0^{\infty} \theta(t)\epsilon_{\mathrm{v}}\lambda e^{-\epsilon_{\mathrm{v}}\lambda\Theta(t)} f_{\mathrm{Gamma}}(\epsilon_{\mathrm{v}}; \alpha_{\mathrm{v}}, \beta_{\mathrm{v}})\, d\epsilon_{\mathrm{v}} = \lambda\theta(t)\bar{\epsilon}_{\mathrm{v}}\left(\frac{\alpha_{\mathrm{v}}}{\alpha_{\mathrm{v}} + \bar{\epsilon}_{\mathrm{v}}\lambda\Theta(t)}\right)^{\alpha_{\mathrm{v}}+1}. \tag{S.30}$$

We calculated two risk ratios, using the instantaneous incidence rates and cumulative attack rates. The instantaneous incidence rate was defined as the probability that a susceptible individual was infected at a given time $t$, $\Pr(\text{infected at time} = t|V = v) = \frac{\mathrm{d}I_{V=v}}{\mathrm{d}t}(t)$. Cumulative attack rates were defined as the probability that a susceptible individual was infected before time $t$, $\Pr(\text{infected before time} = t|V = v) = 1 - \Pr(\text{susceptible at time} = t|V = v) = 1 - S_{V=v}(t)$. $\widehat{\mathrm{VE}}$ was calculated as one minus the infection risk ratio between the vaccinated and unvaccinated populations, using instantaneous incidence rates ($\widehat{\mathrm{VE}}^{\text{instantaneous}}$) or cumulative attack rates ($\widehat{\mathrm{VE}}^{\text{cumulative}}$):

$$\widehat{\mathrm{VE}}^{\text{instantaneous}}(t) = \left(1 - \frac{\frac{\mathrm{d}I_{\mathrm{v}}}{\mathrm{d}t}(t)}{\frac{\mathrm{d}I_{\mathrm{u}}}{\mathrm{d}t}(t)}\right) \times 100\% = \left(1 - \theta(t)\frac{\bar{\epsilon}_{\mathrm{v}}}{\bar{\epsilon}_{\mathrm{u}}}\left[\frac{\left(\frac{\alpha_{\mathrm{v}}}{\alpha_{\mathrm{v}}+\bar{\epsilon}_{\mathrm{v}}\lambda\Theta(t)}\right)^{\alpha_{\mathrm{v}}+1}}{\left(\frac{\alpha_{\mathrm{u}}}{\alpha_{\mathrm{u}}+\bar{\epsilon}_{\mathrm{u}}\lambda t}\right)^{\alpha_{\mathrm{u}}+1}}\right]\right) \times 100\%, \tag{S.31}$$

$$\widehat{\mathrm{VE}}^{\text{cumulative}}(t) = \left(1 - \frac{1 - S_{\mathrm{v}}(t)}{1 - S_{\mathrm{u}}(t)}\right) \times 100\% = \left(1 - \frac{1 - \left(\frac{\alpha_{\mathrm{v}}}{\alpha_{\mathrm{v}}+\bar{\epsilon}_{\mathrm{v}}\lambda\Theta(t)}\right)^{\alpha_{\mathrm{v}}}}{1 - \left(\frac{\alpha_{\mathrm{u}}}{\alpha_{\mathrm{u}}+\bar{\epsilon}_{\mathrm{u}}\lambda t}\right)^{\alpha_{\mathrm{u}}}}\right) \times 100\%. \tag{S.32}$$

###### S.1.2.1 Main text models of estimated VE

We assumed that vaccine direct effects were constant over time, such that $\theta(t) = \theta_0$ and $\Theta(t) = \theta_0 t$ (Supplemental Equations S.12 and S.13 respectively). We substituted the expressions for constant vaccine direct effects into Supplemental Equations S.31 and S.32 to derive the main text VE estimates:

$$\widehat{\mathrm{VE}}^{\text{instantaneous}}(t) = \left(1 - \theta_0\frac{\bar{\epsilon}_{\mathrm{v}}}{\bar{\epsilon}_{\mathrm{u}}}\left[\frac{\left(\frac{\alpha_{\mathrm{v}}}{\alpha_{\mathrm{v}}+\bar{\epsilon}_{\mathrm{v}}\lambda\theta_0 t}\right)^{\alpha_{\mathrm{v}}+1}}{\left(\frac{\alpha_{\mathrm{u}}}{\alpha_{\mathrm{u}}+\bar{\epsilon}_{\mathrm{u}}\lambda t}\right)^{\alpha_{\mathrm{u}}+1}}\right]\right) \times 100\%, \tag{S.33}$$

$$\widehat{\mathrm{VE}}^{\text{cumulative}}(t) = \left(1 - \frac{1 - \left(\frac{\alpha_{\mathrm{v}}}{\alpha_{\mathrm{v}}+\bar{\epsilon}_{\mathrm{v}}\lambda\theta_0 t}\right)^{\alpha_{\mathrm{v}}}}{1 - \left(\frac{\alpha_{\mathrm{u}}}{\alpha_{\mathrm{u}}+\bar{\epsilon}_{\mathrm{u}}\lambda t}\right)^{\alpha_{\mathrm{u}}}}\right) \times 100\%. \tag{S.34}$$

###### S.1.2.2 VE estimated from logistic regression models

We first calculated the expected cumulative counts of test-positive and test-negative infections. For the test-positive infections, we calculated the cumulative attack rates, $1 - S_{V=v}(t)$, using Supplemental Equations S.21 and S.22. We assumed that test-negative infections do not generate protective immunity against subsequent test-negative infections, such that the cumulative test-negative infection hazard is $\lambda_0 t$. We assumed that the probabilities of seeking care ($\mu$) and developing symptoms ($\pi$) do not depend on vaccination status or infection etiology, $\mu_{\text{v}} = \mu_{\text{u}} = \pi_1 = \pi_0$, so that there is no additional confounding in VE estimates. Starting from Supplemental Equations S.5–S.8, we derived the cumulative counts of test-positive and test-negative infections using the parameters in Supplemental Table S.2:

$$X_{11}^{\text{cumulative}}(t) = N\mu_{\text{v}}\pi_1 c\,[1 - S_{\text{v}}(t)] = N\mu_{\text{v}}\pi_1 c\left[1 - \left(\frac{\alpha_{\text{v}}}{\alpha_{\text{v}} + \bar{\epsilon}_{\text{v}}\lambda\theta_0 t}\right)^{\alpha_{\text{v}}}\right], \tag{S.35}$$

$$X_{01}^{\text{cumulative}}(t) = N\mu_{\text{v}}\pi_1(1-c)\,[1 - S_{\text{u}}(t)] = N\mu_{\text{u}}\pi_1(1-c)\left[1 - \left(\frac{\alpha_{\text{u}}}{\alpha_{\text{u}} + \bar{\epsilon}_{\text{u}}\lambda t}\right)^{\alpha_{\text{u}}}\right], \tag{S.36}$$

$$X_{10}^{\text{cumulative}}(t) = N\mu_{\text{v}}\pi_0 c\lambda_0 t\,, \tag{S.37}$$

$$X_{00}^{\text{cumulative}}(t) = N\mu_{\text{u}}\pi_0(1-c)\lambda_0 t\,. \tag{S.38}$$

| Parameter | Description |
|---|---|
| $\lambda$ | Constant exogenous infection hazard of test-positive pathogens |
| $\lambda_0$ | Constant exogenous infection hazard of test-negative pathogens |
| $\theta_0$ | Vaccine direct effects |
| $\bar{\epsilon}_{V=v}$ | Pre-vaccination susceptibility distribution mean for vaccination status $V = v$ |
| $\alpha_{V=v}$ | Pre-vaccination susceptibility distribution shape parameter for vaccination status $V = v$ |
| $\pi_{I=i}$ | Probability of symptoms for infection of cause $I = i$ or $\Pr(S = 1 \mid I = i)$ |
| $\mu_{V=v}$ | Probability of seeking medical care for vaccination status $V = v$ or $\Pr(H = 1 \mid V = v)$ |
| $c$ | Vaccination coverage or $\Pr(V = 1)$ |
| $N$ | Total population size |

**Table S.2:** Parameters for cumulative expected case and control count expressions.

To estimate VE with logistic regression, we generated synthetic datasets by calculating the number of infections in two-week intervals from the cumulative counts. VE was estimated using unconditional logistic regression ($\widehat{\text{VE}}^{\text{unconditional}}$) and conditional logistic regression ($\widehat{\text{VE}}^{\text{conditional}}$). We adjusted for time in both regression methods: with unconditional regression, time is a categorical covariate representing two-week intervals, and with conditional regression, data is matched by time in two-week intervals.

###### S.1.2.3 VE estimated under a cohort design

We defined estimated VE under a cohort study, $\widehat{\text{VE}}^{\text{cohort}}$, as one minus the ratio of average instantaneous incidence rates between the vaccinated and unvaccinated populations, similar to the $\text{VE}_{\text{f}}$ estimate from Smith et al. 1984[3]. The average instantaneous incidence rate equals the cumulative attack rate divided by the expected number of person-time at risk (PTAR). Expected PTAR at time $t$ in the unvaccinated and vaccinated populations is calculated by integrating the susceptible fractions, $S_{\text{u}}$ and $S_{\text{v}}$ (Supplemental Equations S.21 and S.22), from 0 to $t$:

$$\mathbb{E}[\text{PTAR}_{\text{u}}](t) = \int_0^t S_{\text{u}}(s)ds\,, \tag{S.39}$$

$$\mathbb{E}[\text{PTAR}_{\text{v}}](t) = \int_0^t S_{\text{v}}(s)ds\,. \tag{S.40}$$

Cumulative attack rates equal $1 - S_{\mathrm{u}}(t)$ and $1 - S_{\mathrm{v}}(t)$, and $\widehat{\mathrm{VE}}^{\mathrm{cohort}}$ is

$$\widehat{\mathrm{VE}}^{\mathrm{cohort}}(t) = \left(1 - \frac{\frac{1-S_{\mathrm{v}}(t)}{\mathbb{E}[\mathrm{PTAR_v}](t)}}{\frac{1-S_{\mathrm{u}}(t)}{\mathbb{E}[\mathrm{PTAR_u}](t)}}\right) \times 100\% \tag{S.41}$$

###### S.1.2.4 Simplified model of estimated VE with waning true vaccine protection

For supplementary analyses with waning vaccine protection, we simplified VE estimates by assuming that pre-vaccination susceptibility is not continuously distributed. Instead, all vaccinated or unvaccinated people have homogeneous pre-vaccination susceptibility, i.e., $\bar{\epsilon}_{\mathrm{v}}$ and $\bar{\epsilon}_{\mathrm{u}}$ are point densities and constant values. Instantaneous and total vaccine direct effects with waning vaccine protection are the same as described in Supplemental Equations S.12 and S.13. We again calculated how vaccinated and unvaccinated susceptible fractions change over time:

$$\frac{\mathrm{d}S_{\mathrm{u}}}{\mathrm{d}t}(t) = -\bar{\epsilon}_{\mathrm{u}}\lambda S_{\mathrm{u}}(t)\,, \tag{S.42}$$

$$\frac{\mathrm{d}S_{\mathrm{v}}}{\mathrm{d}t}(t) = -\theta(t)\bar{\epsilon}_{\mathrm{v}}\lambda S_{\mathrm{v}}(t)\,. \tag{S.43}$$

We assumed that all individuals begin susceptible and solved for proportion of the two populations susceptible at a given time:

$$S_{\mathrm{u}}(t) = e^{-\lambda\bar{\epsilon}_{\mathrm{u}}t}\,, \tag{S.44}$$

$$S_{\mathrm{v}}(t) = e^{-\lambda\bar{\epsilon}_{\mathrm{v}}\Theta(t)}\,. \tag{S.45}$$

The instantaneous incidence rates in the vaccinated and unvaccinated populations are

$$\frac{\mathrm{d}I_{\mathrm{u}}}{\mathrm{d}t}(t) = \bar{\epsilon}_{\mathrm{u}}\lambda S_{\mathrm{u}}(t) = \bar{\epsilon}_{\mathrm{u}}\lambda e^{-\bar{\epsilon}_{\mathrm{u}}\lambda t}\,, \tag{S.46}$$

$$\frac{\mathrm{d}I_{\mathrm{v}}}{\mathrm{d}t}(t) = \theta(t)\bar{\epsilon}_{\mathrm{v}}\lambda S_{\mathrm{v}}(t) = \theta(t)\bar{\epsilon}_{\mathrm{v}}\lambda e^{-\bar{\epsilon}_{\mathrm{v}}\lambda\Theta(t)}\,. \tag{S.47}$$

Therefore, the VE estimates with waning vaccine protection are

$$\widehat{\mathrm{VE}}^{\mathrm{instantaneous}}(t) = \left(1 - \theta(t)\frac{\bar{\epsilon}_{\mathrm{v}}}{\bar{\epsilon}_{\mathrm{u}}}e^{-\lambda[\bar{\epsilon}_{\mathrm{v}}\Theta(t) - \bar{\epsilon}_{\mathrm{u}}t]}\right) \times 100\%\,, \tag{S.48}$$

$$\widehat{\mathrm{VE}}^{\mathrm{cumulative}}(t) = \left(1 - \frac{1 - e^{-\bar{\epsilon}_{\mathrm{v}}\lambda\Theta(t)}}{1 - e^{-\bar{\epsilon}_{\mathrm{u}}\lambda t}}\right) \times 100\%\,. \tag{S.49}$$

We substituted Supplemental Equation S.45 into regression (Supplemental Equation S.35) and cohort (Supplemental Equation S.41) $\widehat{\mathrm{VE}}$ estimates to perform $\widehat{\mathrm{VE}}$ comparisons with waning vaccine protection.

##### S.1.3 VE estimate comparisons

To compare $\widehat{\mathrm{VE}}$ from different estimation methods, we calculated differences in $\widehat{\mathrm{VE}}^{\mathrm{instantaneous}}$, $\widehat{\mathrm{VE}}^{\mathrm{unconditional}}$, $\widehat{\mathrm{VE}}^{\mathrm{conditional}}$, and $\widehat{\mathrm{VE}}^{\mathrm{cohort}}$ estimates relative to $\widehat{\mathrm{VE}}^{\mathrm{cumulative}}$ estimates. We reported the average and 95% interval of the differences. The parameter values used for each set of comparisons are in Supplemental Table S.3, and final VE estimates were calculated after 196 days to ensure an integral number of two-week intervals.

| Parameter | Value(s) for main results section 1 comparisons | Value(s) for main results section 2 comparisons | Value(s) for supplemental waning results comparisons |
|---|---|---|---|
| $\lambda$ | 0.005 | 0.0015 | 0.005 |
| $\lambda_0$ | 0.015 | 0.0045 | 0.015 |
| $\theta_0$ | $\{0.1, 0.2, ..., 0.9\}$ | $\{0.01, 0.02, ..., 0.99\}$ | $\{0.1, 0.2, ..., 0.9\}$ |
| $\eta$ | NA | NA | $\{30, 180, 360, 1440\}$ |
| $\bar{\epsilon}_\text{v}$ | $\{0.1, 0.2, ..., 1.0\}$ | 1 | $\{0.1, 0.2, ..., 1.0\}$ |
| $\bar{\epsilon}_\text{u}$ | $\{0.1, 0.2, ..., 1.0\}$ | 1 | $\{0.1, 0.2, ..., 1.0\}$ |
| $\alpha_\text{v}$ | 20 | $\{0.2, 2, 20\}$ | NA |
| $\alpha_\text{u}$ | 20 | $\{0.2, 2, 20\}$ | NA |
| $\pi_1$ | 1 | 1 | 1 |
| $\pi_0$ | 1 | 1 | 1 |
| $\mu_\text{v}$ | 1 | 1 | 1 |
| $\mu_\text{u}$ | 1 | 1 | 1 |
| $c$ | 0.5 | 0.5 | 0.5 |
| $N$ | $5 \times 10^3$ | $5 \times 10^3$ | $5 \times 10^3$ |

**Table S.3:** Parameters for cumulative expected case and control count expressions. NA = not applicable.

##### S.1.4 Simulation methods

We developed an individual-based model of seasonal epidemics in a partially vaccinated population. The synthetic population is composed of five million individuals, and we did not model any births or deaths. Half of the population is initially vaccinated, and each individual's pre-vaccination susceptibility is a random sample from a gamma distribution parameterized based on the individual's initial vaccination status. The vaccinated and unvaccinated populations start with the same mean pre-vaccination susceptibilities ($\bar{\epsilon}_\text{u} = \bar{\epsilon}_\text{v} = 1$), but the unvaccinated population is more heterogeneous than the vaccinated population ($\alpha_\text{v} = 10$, $\alpha_\text{u} = 1$).

The infection hazard $\lambda(t)$ is exogenous and equal for all individuals. It is seasonally forced to generate annual epidemics, $\lambda(t) = \beta_0 [1 + \cos(2\pi(t + 0.5))]$. In most simulated years, the infection hazard generates epidemics with an average cumulative attack rate of about 25% ($\beta_0 = 1$). In the fourth year of the simulation, the infection hazard increases to simulate a larger-than-normal epidemic with a cumulative attack rate of approximately 60% ($\beta_0 = 4$).

An individual's risk of infection is modified by their infection- and vaccine-derived immune protection. Infections generate initially perfect immune protection that wanes exponentially with a four-year half life[4] to the individual's pre-vaccination susceptibility level. Vaccination generates 60% leaky protection (see Supplemental Figures S.9 and S.10 for simulations with higher and lower true vaccine protection), and vaccine-derived immune protection is constant over time. At the start of each simulated year, the population is re-vaccinated. There is an 85% probability that a currently vaccinated individual is re-vaccinated in the next year[5,6]. Using the vaccinated-to-vaccinated transition probability, we calculated the corresponding unvaccinated-to-vaccinated transition probability to maintain a vaccination coverage of 50% each year. All vaccine-derived protection is lost when individuals switches from vaccinated to unvaccinated.

We simulated infections using an accept-reject algorithm, following the guidelines in Wu et al[7]. During a single time step (0.005 years), we calculated an individual's force of infection using the current exogenous infection hazard, their current susceptibility, and any vaccine-derived protection. For each susceptible individual, we sampled the time until their next infection from an exponential distribution using the force of infection as the rate parameter. If the sampled infection time occurs before the end of the current time step, the individual is infected. Otherwise, the individual remained susceptible for the rest of the time step.

To estimate VE, we first estimated counts of test-negative infections. The test-negative infection hazard follows the same seasonal forcing as the test-positive infection hazard but with a higher cumulative attack rate (approximately 75%). To ensure conditional regression estimates converge, we randomly sampled 0.15% of each year's infection data and used the smaller datasets, approximately 6000–9000 total test-positive and test-negative infections per year, to estimate $\widehat{\text{VE}}^{\text{cumulative}}$, $\widehat{\text{VE}}^{\text{cohort}}$, $\widehat{\text{VE}}^{\text{unconditional}}$, and $\widehat{\text{VE}}^{\text{conditional}}$ estimates. $\widehat{\text{VE}}^{\text{cumulative}}$ is estimated as one minus the ratio of the annual cumulative attack rates between the vaccinated and unvaccinated populations. $\widehat{\text{VE}}^{\text{cohort}}$ is estimated as one minus the ratio of the annual average instantaneous incidence rates between the vaccinated and unvaccinated populations. The average instantaneous incidence rate is calculated by dividing the total number of infections by the total PTAR across individuals. Each year, infected individuals' PTAR equals their infection time, whereas uninfected individuals' PTAR equals one. For regression-based $\widehat{\text{VE}}$ estimates, the sampled test-positive and test-negative infections for each year are binned into two-week intervals. We then estimated $\widehat{\text{VE}}$ using conditional and unconditional logistic regression, adjusting or matching for time. As in our other $\widehat{\text{VE}}$ estimates, we assumed that there is no unmeasured confounding from the probability of developing symptoms or seeking medical care.

We compared $\widehat{\text{VE}}$ from cohort and logistic regression methods to $\widehat{\text{VE}}^{\text{cumulative}}$ (Supplemental Figure S.11). We did not calculate final $\widehat{\text{VE}}^{\text{instantaneous}}$ estimates because they is more sensitive to differential depletion of susceptibles[1] and are not commonly estimated in real-world observational studies. We again reported the mean and 95% interval of the differences between $\widehat{\text{VE}}^{\text{cumulative}}$ and other $\widehat{\text{VE}}$ estimates.

##### S.1.5 Software used

Multiyear simulations were implemented in Julia 1.10.2, and all $\widehat{\text{VE}}$ analyses were performed in R 4.3.3. The `survival` R package (version 3.8.6) was used to perform conditional logistic regression. All software is available at `https://github.com/cobeylab/heterogeneity_in_pre-vax_immunity_contributes_to_ve_variability`.

#### S.2 Supplemental results

##### S.2.1 Derivation of main text equation 2

To simplify the derivations, we evaluated the limits of the $\widehat{\text{VE}}$ estimates divided by 100%.

###### S.2.1.1 Limit of estimated VE using cumulative attack rates as time approaches zero

Starting with Supplemental Equation S.34, let $\text{A} = \theta_0 \bar{\epsilon}_{\text{v}} \lambda$ and $\text{B} = \bar{\epsilon}_{\text{u}} \lambda$:

$$\widehat{\text{VE}}^{\text{cumulative}}(t) = 1 - \frac{1 - \left(\frac{\alpha_{\text{v}}}{\alpha_{\text{v}} + \text{A}t}\right)^{\alpha_{\text{v}}}}{1 - \left(\frac{\alpha_{\text{u}}}{\alpha_{\text{u}} + \text{B}t}\right)^{\alpha_{\text{u}}}}. \tag{S.50}$$

Taking the limit as time approaches zero,

$$\lim_{t\to 0}\widehat{\mathrm{VE}}^{\mathrm{cumulative}}(t) = 1 - \lim_{t\to 0}\frac{1-\left(\frac{\alpha_{\mathrm{v}}}{\alpha_{\mathrm{v}}+\mathrm{A}t}\right)^{\alpha_{\mathrm{v}}}}{1-\left(\frac{\alpha_{\mathrm{u}}}{\alpha_{\mathrm{u}}+\mathrm{B}t}\right)^{\alpha_{\mathrm{u}}}}$$
$$= 1 - \frac{0}{0}\,. \tag{S.51}$$

To apply L'Hôpital's Rule, we first evaluated the following derivative with respect to time. The derivative represents a generalized version of the numerator and denominator of Supplemental Equation S.50:

$$\begin{aligned}\frac{\mathrm{d}}{\mathrm{d}t}\,1-\left(\frac{a}{a+bt}\right)^{a} &= 0-\frac{\mathrm{d}}{\mathrm{d}t}\left(\frac{a}{a+bt}\right)^{a}\\ &= -a\left(\frac{a}{a+bt}\right)^{a-1}\frac{\mathrm{d}}{\mathrm{d}t}\left(\frac{a}{a+bt}\right)\\ &= -a^2\left(\frac{a}{a+bt}\right)^{a-1}\frac{\mathrm{d}}{\mathrm{d}t}\,(a+bt)^{-1}\\ &= a^2\left(\frac{a}{a+bt}\right)^{a-1}(a+bt)^{-2}\,\frac{\mathrm{d}}{\mathrm{d}t}\,(a+bt)\\ &= b\left(\frac{a}{a+bt}\right)^{a+1}\,.\end{aligned} \tag{S.52}$$

Using the generalized derivation as a guide, we calculated the derivatives with respect to time of the numerator and denominator of Supplemental Equation S.50:

$$\frac{\mathrm{d}}{\mathrm{d}t}\,1-\left(\frac{\alpha_{\mathrm{v}}}{\alpha_{\mathrm{v}}+\mathrm{A}t}\right)^{\alpha_{\mathrm{v}}} = A\left(\frac{\alpha_{\mathrm{v}}}{\alpha_{\mathrm{v}}+\mathrm{A}t}\right)^{\alpha_{\mathrm{v}}+1}, \tag{S.53}$$
$$\frac{\mathrm{d}}{\mathrm{d}t}\,1-\left(\frac{\alpha_{\mathrm{u}}}{\alpha_{\mathrm{u}}+\mathrm{B}t}\right)^{\alpha_{\mathrm{u}}} = B\left(\frac{\alpha_{\mathrm{u}}}{\alpha_{\mathrm{u}}+\mathrm{B}t}\right)^{\alpha_{\mathrm{u}}+1}. \tag{S.54}$$

We applied L'Hôpital's Rule, and the limit becomes

$$\begin{aligned}\lim_{t\to 0}\widehat{\mathrm{VE}}^{\mathrm{cumulative}}(t) &= 1-\lim_{t\to 0}\frac{A\left(\frac{\alpha_{\mathrm{v}}}{\alpha_{\mathrm{v}}+\mathrm{A}t}\right)^{\alpha_{\mathrm{v}}+1}}{B\left(\frac{\alpha_{\mathrm{u}}}{\alpha_{\mathrm{u}}+\mathrm{B}t}\right)^{\alpha_{\mathrm{u}}+1}}\\ &= 1-\frac{A}{B}\\ &= 1-\frac{\lambda\bar{\epsilon}_{\mathrm{v}}\theta_0}{\lambda\bar{\epsilon}_{\mathrm{u}}}\\ &= 1-\theta_0\,\frac{\bar{\epsilon}_{\mathrm{v}}}{\bar{\epsilon}_{\mathrm{u}}}\,.\end{aligned} \tag{S.55}$$

###### S.2.1.2 Limit of estimated VE using instantaneous incidence rates as time approaches zero

From Supplemental Equation S.33,

$$\begin{aligned}
\lim_{t\to 0}\widehat{\mathrm{VE}}^{\text{instantaneous}}(t) &= \lim_{t\to 0} 1-\theta_0\frac{\bar{\epsilon}_{\mathrm{v}}}{\bar{\epsilon}_{\mathrm{u}}}\left[\frac{\left(\frac{\alpha_{\mathrm{v}}}{\alpha_{\mathrm{v}}+\bar{\epsilon}_{\mathrm{v}}\lambda\theta_0 t}\right)^{\alpha_{\mathrm{v}}+1}}{\left(\frac{\alpha_{\mathrm{u}}}{\alpha_{\mathrm{u}}+\bar{\epsilon}_{\mathrm{u}}\lambda t}\right)^{\alpha_{\mathrm{u}}+1}}\right] \\
&= 1-\theta_0\frac{\bar{\epsilon}_{\mathrm{v}}}{\bar{\epsilon}_{\mathrm{u}}}\lim_{t\to 0}\left[\frac{\left(\frac{\alpha_{\mathrm{v}}}{\alpha_{\mathrm{v}}+\bar{\epsilon}_{\mathrm{v}}\lambda\theta_0 t}\right)^{\alpha_{\mathrm{v}}+1}}{\left(\frac{\alpha_{\mathrm{u}}}{\alpha_{\mathrm{u}}+\bar{\epsilon}_{\mathrm{u}}\lambda t}\right)^{\alpha_{\mathrm{u}}+1}}\right] \\
&= 1-\theta_0\frac{\bar{\epsilon}_{\mathrm{v}}}{\bar{\epsilon}_{\mathrm{u}}}\,. \qquad \text{(S.56)}
\end{aligned}$$

###### S.2.1.3 Limit of estimated VE under a cohort design as time approaches zero

Starting from Supplemental Equation S.41, , let $\mathrm{A}=\theta_0\bar{\epsilon}_{\mathrm{v}}\lambda$ and $\mathrm{B}=\bar{\epsilon}_{\mathrm{u}}\lambda$:

$$\begin{aligned}
\lim_{t\to 0}\widehat{\mathrm{VE}}^{\text{cohort}}(t) &= \lim_{t\to 0} 1-\frac{\frac{1-S_{\mathrm{v}}(t)}{\mathbb{E}[\mathrm{PTAR}_{\mathrm{v}}](t)}}{\frac{1-S_{\mathrm{u}}(t)}{\mathbb{E}[\mathrm{PTAR}_{\mathrm{u}}](t)}} \\
&= 1-\frac{\lim_{t\to 0}\frac{1-\left(\frac{\alpha_{\mathrm{v}}}{\alpha_{\mathrm{v}}+\mathrm{A}t}\right)^{\alpha_{\mathrm{v}}}}{\int_0^t\left(\frac{\alpha_{\mathrm{v}}}{\alpha_{\mathrm{v}}+\mathrm{A}s}\right)^{\alpha_{\mathrm{v}}}ds}}{\lim_{t\to 0}\frac{1-\left(\frac{\alpha_{\mathrm{u}}}{\alpha_{\mathrm{u}}+\mathrm{B}t}\right)^{\alpha_{\mathrm{u}}}}{\int_0^t\left(\frac{\alpha_{\mathrm{u}}}{\alpha_{\mathrm{u}}+\mathrm{B}s}\right)^{\alpha_{\mathrm{u}}}ds}} \qquad \text{(S.57)}
\end{aligned}$$

To evaluate the limit, we started with a generalized version of the average instantaneous incidence rates in the numerator and denominator,

$$\lim_{t\to 0}\frac{1-\left(\frac{a}{a+bt}\right)^a}{\int_0^t\left(\frac{a}{a+bs}\right)^a ds}=\frac{0}{0}\,. \qquad \text{(S.58)}$$

To apply L'Hôpital's Rule, we calculated the derivative with respect to time. The numerator's derivative is the same as Supplemental Equation S.52. The denominator's derivative is

$$\frac{\mathrm{d}}{\mathrm{d}t}\int_0^t\left(\frac{a}{a+bs}\right)^a ds=\left(\frac{a}{a+bt}\right)^a\,. \qquad \text{(S.59)}$$

The generalized limit therefore becomes

$$\begin{aligned}
\lim_{t\to 0}\frac{1-\left(\frac{a}{a+bt}\right)^a}{\int_0^t\left(\frac{a}{a+bs}\right)^a ds} &= \lim_{t\to 0}\frac{b\left(\frac{a}{a+bt}\right)^{a+1}}{\left(\frac{a}{a+bt}\right)^a} \\
&= \lim_{t\to 0} b\left(\frac{a}{a+bt}\right) \\
&= b \qquad \text{(S.60)}
\end{aligned}$$

Using the generalized derivation as a guide, we solved the original limit:

$$\begin{aligned}
\lim_{t\to 0} \widehat{\mathrm{VE}}^{\text{cohort}}(t) &= 1 - \frac{\lim_{t\to 0} \frac{1-\left(\frac{\alpha_{\mathrm{v}}}{\alpha_{\mathrm{v}}+\mathrm{A}t}\right)^{\alpha_{\mathrm{v}}}}{\int_0^t \left(\frac{\alpha_{\mathrm{v}}}{\alpha_{\mathrm{v}}+\mathrm{A}s}\right)^{\alpha_{\mathrm{v}}} ds}}{\lim_{t\to 0} \frac{1-\left(\frac{\alpha_{\mathrm{u}}}{\alpha_{\mathrm{u}}+\mathrm{B}t}\right)^{\alpha_{\mathrm{u}}}}{\int_0^t \left(\frac{\alpha_{\mathrm{u}}}{\alpha_{\mathrm{u}}+\mathrm{B}s}\right)^{\alpha_{\mathrm{u}}} ds}} \\
&= 1 - \frac{A}{B} \\
&= 1 - \frac{\lambda \bar{\epsilon}_{\mathrm{v}} \theta_0}{\lambda \bar{\epsilon}_{\mathrm{u}}} \\
&= 1 - \theta_0 \, \frac{\bar{\epsilon}_{\mathrm{v}}}{\bar{\epsilon}_{\mathrm{u}}} \, .
\end{aligned} \tag{S.61}$$

##### S.2.2 Estimated VE can become negative when mean pre-vaccination susceptibilities differ and true vaccine protection wanes

Selecting a reference value to compare VE estimates against is challenging when vaccine protection wanes. If VE is estimated with instantaneous incidence rates, then we would compare $\widehat{\mathrm{VE}}^{\text{instantaneous}}$ to the vaccine's instantaneous level of protection across time, $\theta(t)$, as prior models have done[8]. However, when VE is estimated with cumulative attack rates, it reflects the total protection of a vaccine over an entire time window rather than the instantaneous level of protection at any given time. One way to estimate cumulative vaccine protection is with the ratio of the total infection hazard between the vaccinated and unvaccinated populations across some time window (Supplemental Figure S.1). The total infection hazard is found by integrating the product of the exogenous infection hazard, pre-vaccination susceptibility, and vaccine protection (if vaccinated),

$$\frac{\int_0^t \lambda\, \bar{\epsilon}_{\mathrm{v}}\, \theta(s)\, ds}{\int_0^t \lambda\, \bar{\epsilon}_{\mathrm{u}}\, ds} = \left(\frac{\bar{\epsilon}_{\mathrm{v}}}{\bar{\epsilon}_{\mathrm{u}}}\right) \times \left(\frac{\int_0^t \theta(s)\, ds}{t}\right) . \tag{S.62}$$

The total hazard ratio reduces to the average protection of the vaccine through time $t$ scaled by the mean pre-vaccination susceptibility ratio. If mean pre-vaccination susceptibility were equal between the vaccinated and unvaccinated populations, the right-hand side of Supplemental Equation S.62 simplifies to the vaccine's average protection. We therefore evaluated the difference between $\widehat{\mathrm{VE}}$ and average true vaccine protection through time $t$. As in the main text, we estimated VE using cumulative attack rates ($\widehat{\mathrm{VE}}^{\text{cumulative}}$), instantaneous incidence rates ($\widehat{\mathrm{VE}}^{\text{instantaneous}}$), average instantaneous incidence rates ($\widehat{\mathrm{VE}}^{\text{cohort}}$), and logistic regression ($\widehat{\mathrm{VE}}^{\text{unconditional}}$ and $\widehat{\mathrm{VE}}^{\text{conditional}}$). Unless otherwise noted, $\widehat{\mathrm{VE}}$ refers to $\widehat{\mathrm{VE}}^{\text{cumulative}}$.

When vaccine protection wanes and mean pre-vaccination susceptibility is equal between the vaccinated and unvaccinated populations, $\widehat{\mathrm{VE}}$ is always less than the average vaccine protection[8]. The difference between $\widehat{\mathrm{VE}}$ and the vaccine's average protection generally shrinks as vaccine protection wanes more quickly (Supplemental Figure S.2). However, when starting vaccine protection is high, $\widehat{\mathrm{VE}}$ differs most from average vaccine protection at an intermediate waning rate. This behavior follows previous results: At a given infection hazard, differential depletion of susceptibles affects $\widehat{\mathrm{VE}}$ most when vaccines generate intermediate protection[1]. When vaccine protection is higher or lower than this intermediate level, $\widehat{\mathrm{VE}}$ is closer to true protection (Supplemental Figure S.3 A). Our waning model similarly finds that the difference between $\widehat{\mathrm{VE}}$ and average vaccine protection is sensitive to whether the average protection is close to an intermediate level, which depends on the starting vaccine protection and rate of waning (Supplemental Figure S.3 B and C).

Waning vaccine protection combined with differential mean pre-vaccination susceptibility between vaccinated and unvaccinated populations can cause $\widehat{\mathrm{VE}}$ to be negative. As expected, the entire $\widehat{\mathrm{VE}}$ trajectory

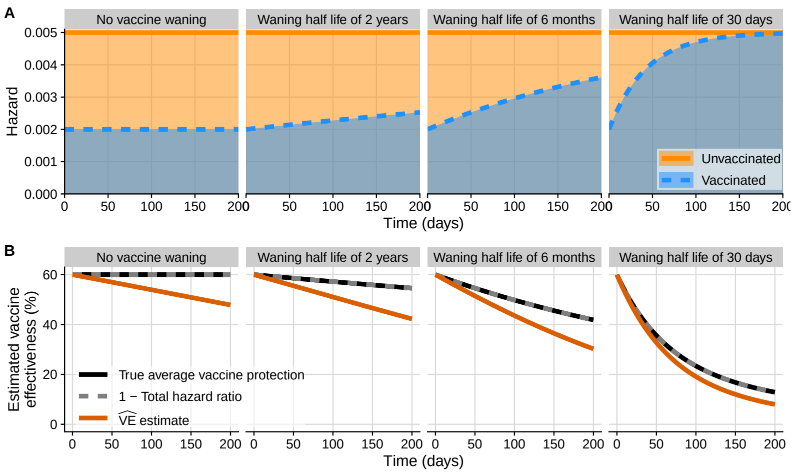

**Figure S.1:** When true vaccine protection wanes, we compared $\widehat{\mathrm{VE}}$ to the vaccine's true average protection. For all subplots, the columns refer to different vaccine protection waning rates, ranging from no waning (left-most column) to rapid waning (right-most column). (A) Time-varying infection hazard in the unvaccinated (orange solid line) and vaccinated (blue dashed line) populations. The shaded regions correspond to the total infection hazard through time $t$. (B) Time-varying $\widehat{\mathrm{VE}}$ (orange lines) compared to the vaccine's average protection (black solid lines). Because mean pre-vaccination susceptibility is equal in vaccinated and unvaccinated people in these scenarios, the total hazard ratio (gray dashed lines) is identical to the average true vaccine protection.

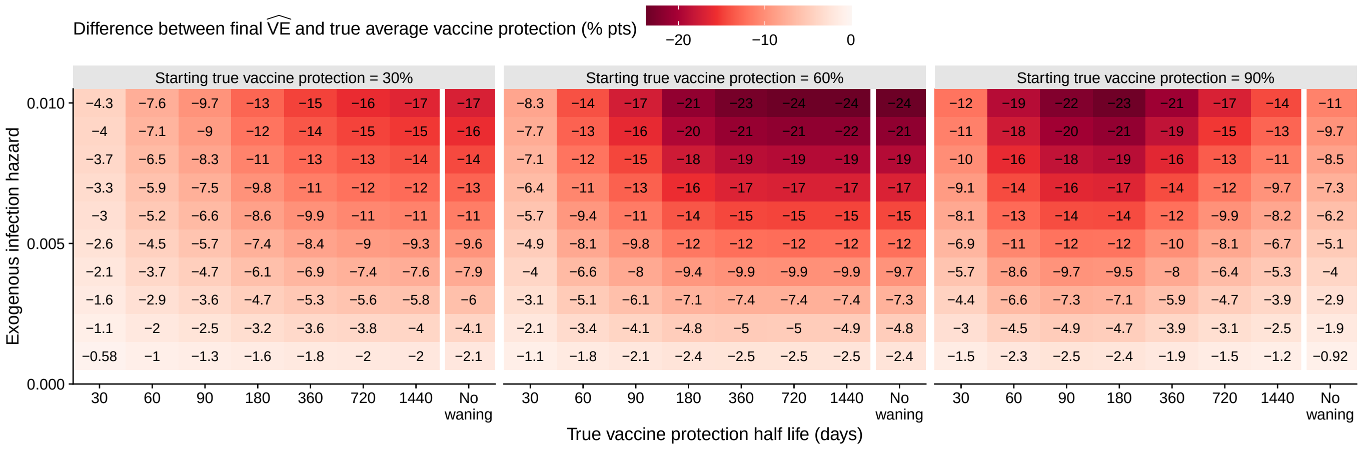

**Figure S.2:** Final $\widehat{\mathrm{VE}}$ is generally closer to true average vaccine protection when vaccine protection wanes quickly. The vaccinated and unvaccinated populations have identical mean pre-vaccination susceptibilities ($\bar{\epsilon}_{\mathrm{v}} = \bar{\epsilon}_{\mathrm{u}} = 1$). Final $\widehat{\mathrm{VE}}$ is calculated after a 200-day epidemic. The conditions under which $\widehat{\mathrm{VE}}$ differs most from average vaccine protection depend on the exogenous infection hazard (y-axis), the vaccine's starting protection (panels) and how quickly its protection wanes (x-axis).

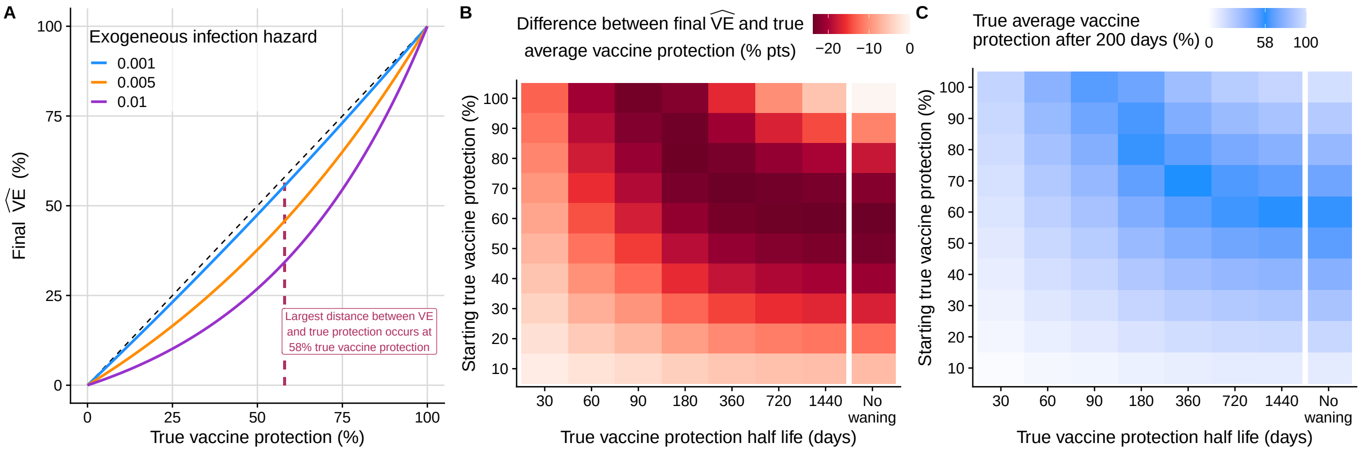

**Figure S.3:** When true vaccine protection wanes, the difference between $\widehat{VE}$ and true average vaccine protection is highest at intermediate levels of average vaccine protection. For all scenarios, final $\widehat{VE}$ is estimated after 200 days, and mean pre-vaccination susceptibility is identical between the vaccinated and unvaccinated populations ($\bar{\epsilon}_v = \bar{\epsilon}_u = 1$). (A) Final $\widehat{VE}$ is further from true vaccine protection when true protection is moderate. This panel is adapted from Lewnard et al. Figure 1 C[1] and assumes that true vaccine protection is constant over time. (B) When starting true vaccine protection is low to moderate (10–60%), the difference between final $\widehat{VE}$ and average vaccine protection is largest (darker red) when waning is slowest or nonexistent. When starting vaccine protection is high (70–100%), the largest difference occurs at an intermediate waning rate. (C) Final average vaccine protection depends on the starting protection and rate of waning. Intermediate true average vaccine protection (darker blue) corresponds to the largest difference between final $\widehat{VE}$ and average vaccine protection (darker red in B). For scenarios in panels B and C, the exogenous infection hazard $\lambda = 0.01$.

shifts higher or lower depending on the mean pre-vaccination susceptibility ratio (Supplemental Figure S.4 top). When vaccine protection decreases over time, $\widehat{VE}$ can start positive and become negative when the unvaccinated population has lower mean pre-vaccination susceptibility compared to the vaccinated population (Supplemental Figure S.4 bottom). As vaccine protection wanes faster or starting vaccine protection increases, more combinations of mean pre-vaccination susceptibility in the two populations result in $\widehat{VE}$ crossing zero and becoming negative (Supplemental Figure S.5).

Differences in mean pre-vaccination susceptibility alongside waning vaccine protection affect $\widehat{VE}$ in similar directions regardless of how it is estimated. Across all waning scenarios, regression-based $\widehat{VE}$ estimates differ from $\widehat{VE}^{cumulative}$ estimates by less than a percentage point on average (95% interval: -2 to 1 percentage-point difference for $\widehat{VE}^{unconditional}$ and $\widehat{VE}^{conditional}$), whereas $\widehat{VE}^{instantaneous}$ are lower than $\widehat{VE}^{cumulative}$ estimates by 13 percentage points on average and vary more (95% interval: -53 to 57 percentage-point difference). Increased variation in $\widehat{VE}$ estimated from instantaneous incidence rates is expected because $\widehat{VE}^{instantaneous}$ is more sensitive to differential depletion of susceptibles[1]. Additionally, comparing $\widehat{VE}^{instantaneous}$ to $\widehat{VE}^{cumulative}$ also reflects differences in the estimators themselves: $\widehat{VE}^{cumulative}$ estimates a vaccine's average protection over an entire time window, whereas $\widehat{VE}^{instantaneous}$ estimates the vaccine's instantaneous protection at a given time. Estimates of $\widehat{VE}^{cohort}$ are 4 percentage points lower than $\widehat{VE}^{cumulative}$ estimates on average and also vary more than regression-based estimates (95% interval: -81 to 12 percentage-point difference). The higher variation in $\widehat{VE}^{cohort}$ estimates is expected because $\widehat{VE}^{cohort}$ is less susceptible to differential depletion of susceptibles[3] and remains closer to starting $\widehat{VE}$ compared to $\widehat{VE}^{cumulative}$.

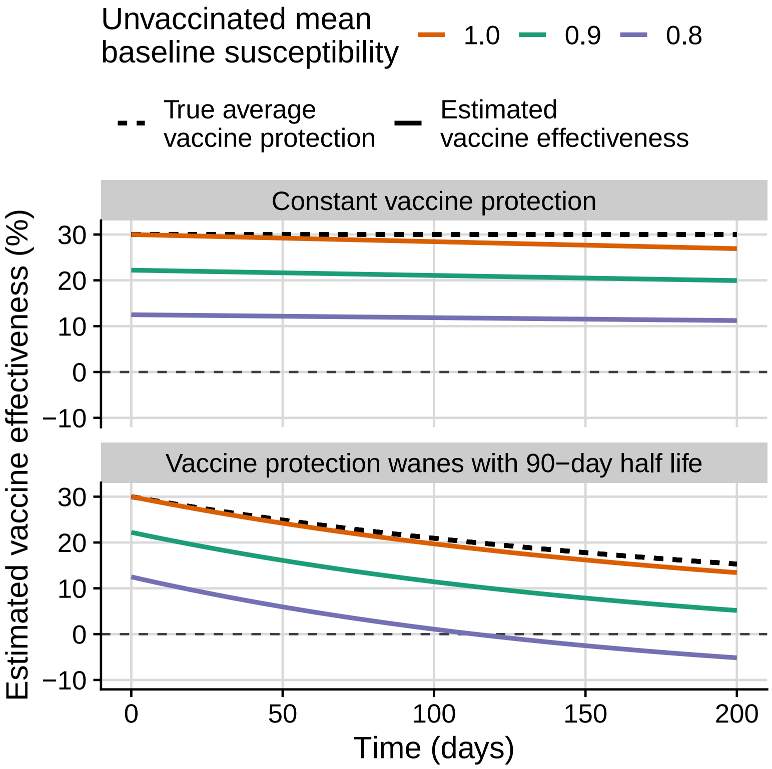

**Figure S.4:** $\widehat{\mathrm{VE}}$ can become negative when vaccine protection wanes over time and the unvaccinated population has lower mean pre-vaccination susceptibility than the vaccinated. Mean pre-vaccination susceptibility in the vaccinated population is constant ($\bar{\epsilon}_{\mathrm{v}} = 1$), while unvaccinated mean pre-vaccination susceptibility varies (solid colored lines). For these example trajectories, the exogenous infection hazard $\lambda = 0.0015$. (Top) When vaccine protection does not wane, the entire $\widehat{\mathrm{VE}}$ trajectory shifts downwards as unvaccinated mean pre-vaccination susceptibility decreases, and all trajectories approach zero. (Bottom) If vaccine protection wanes over time (with a 90 day half life in this case), $\widehat{\mathrm{VE}}$ can start positive and become negative during the epidemic when the unvaccinated population has lower mean pre-vaccination susceptibility (purple line).

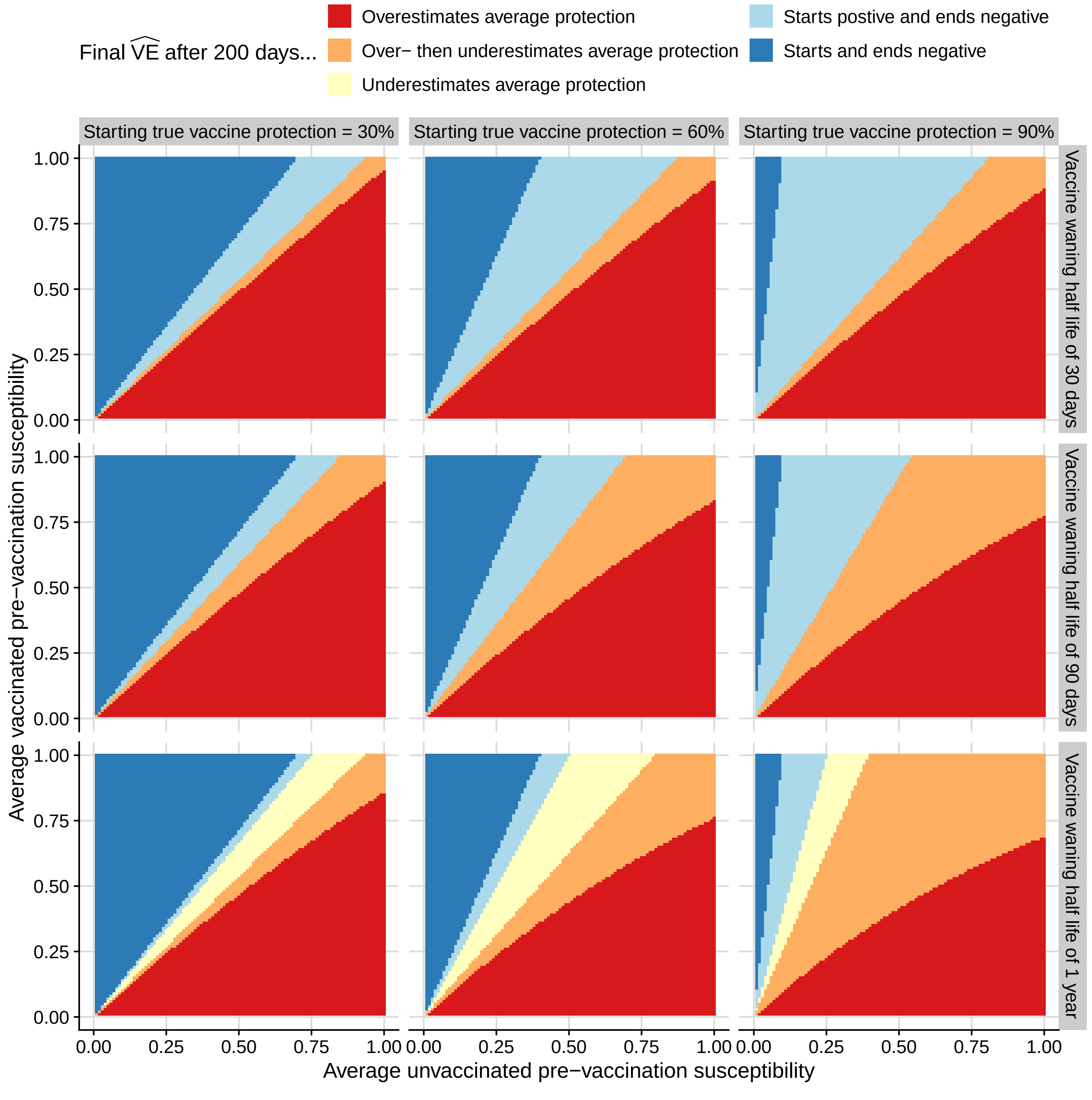

**Figure S.5:** $\widehat{\text{VE}}$ has a higher chance of crossing zero if vaccine-derived protection wanes faster or starts higher. Comparisons of final $\widehat{\text{VE}}$ and average true vaccine protection (red, orange, and yellow) involve $\widehat{\text{VE}}$ that starts and ends positive. Light blue corresponds to scenarios where $\widehat{\text{VE}}$ starts lower than true protection but eventually crosses zero and ends negative. Dark blue corresponds to scenarios where $\widehat{\text{VE}}$ starts and ends negative. In all scenarios, the exogenous infection hazard $\lambda = 0.005$.

### S.3 Supplemental figures

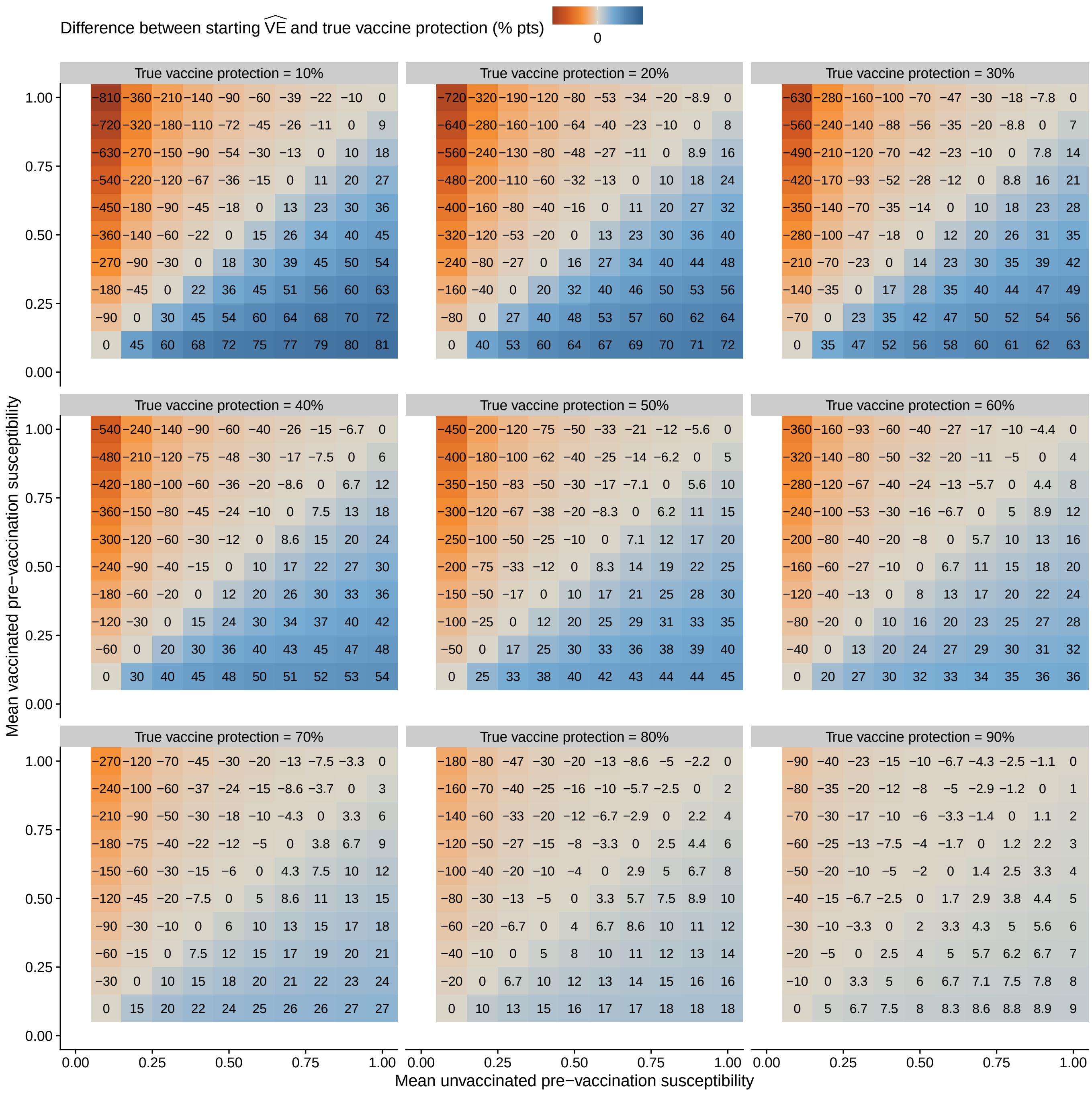

**Figure S.6:** Starting $\widehat{\mathrm{VE}}$ differs from true vaccine protection when vaccinated and unvaccinated populations have different mean pre-vaccination susceptibilities. The vaccinated and unvaccinated distributions of pre-vaccination susceptibility have different means but identical shape parameters corresponding to a highly homogeneous distribution ($\alpha_v = \alpha_u = 20$). Darker blue corresponds to starting $\widehat{\mathrm{VE}}$ that exceeds true vaccine protection and darker orange corresponds to $\widehat{\mathrm{VE}}$ that starts lower than true vaccine protection. Note that for the same mean pre-vaccination susceptibility ratio (i.e., combination of x- and y-axis value), the difference between $\widehat{\mathrm{VE}}$ and true vaccine protection decreases as true protection increases.

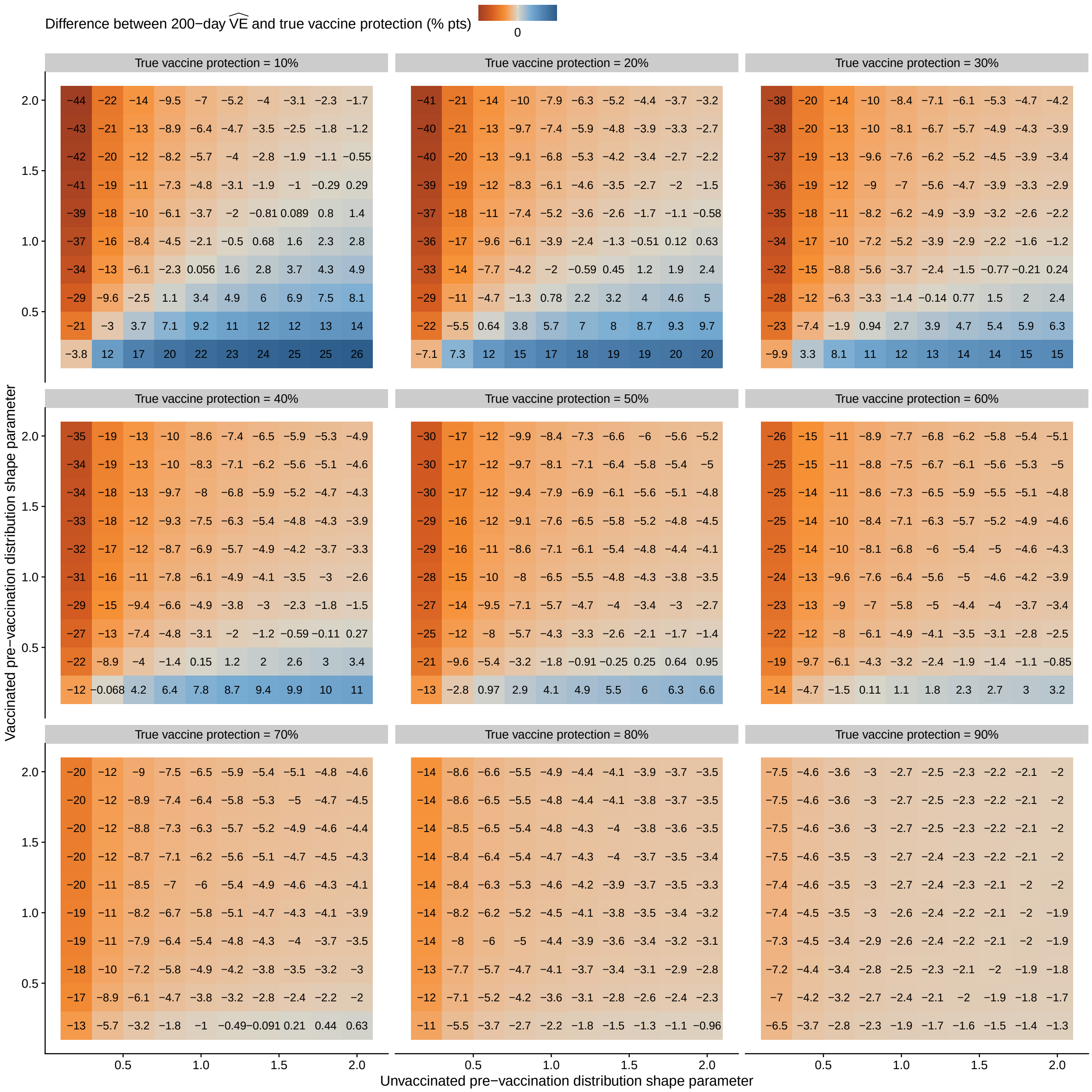

**Figure S.7:** High heterogeneity in pre-vaccination susceptibility causes larger differences between final $\widehat{\text{VE}}$ and true vaccine protection. In all scenarios, mean pre-vaccination susceptibility is identical between vaccinated and unvaccinated people ($\bar{\epsilon}_v = \bar{\epsilon}_u = 1$). Epidemics last for 200 days with a constant exogenous infection hazard $\lambda = 0.0015$. We vary the shape parameters of the vaccinated and unvaccinated pre-vaccination susceptibility distributions from 0.2 (more heterogeneous) to 2 (less heterogeneous). Darker cell colors correspond to larger differences between final $\widehat{\text{VE}}$ and true vaccine protection, with orange corresponding to $\widehat{\text{VE}}$ that is less than true protection and blue corresponding to $\widehat{\text{VE}}$ that exceeds true protection.

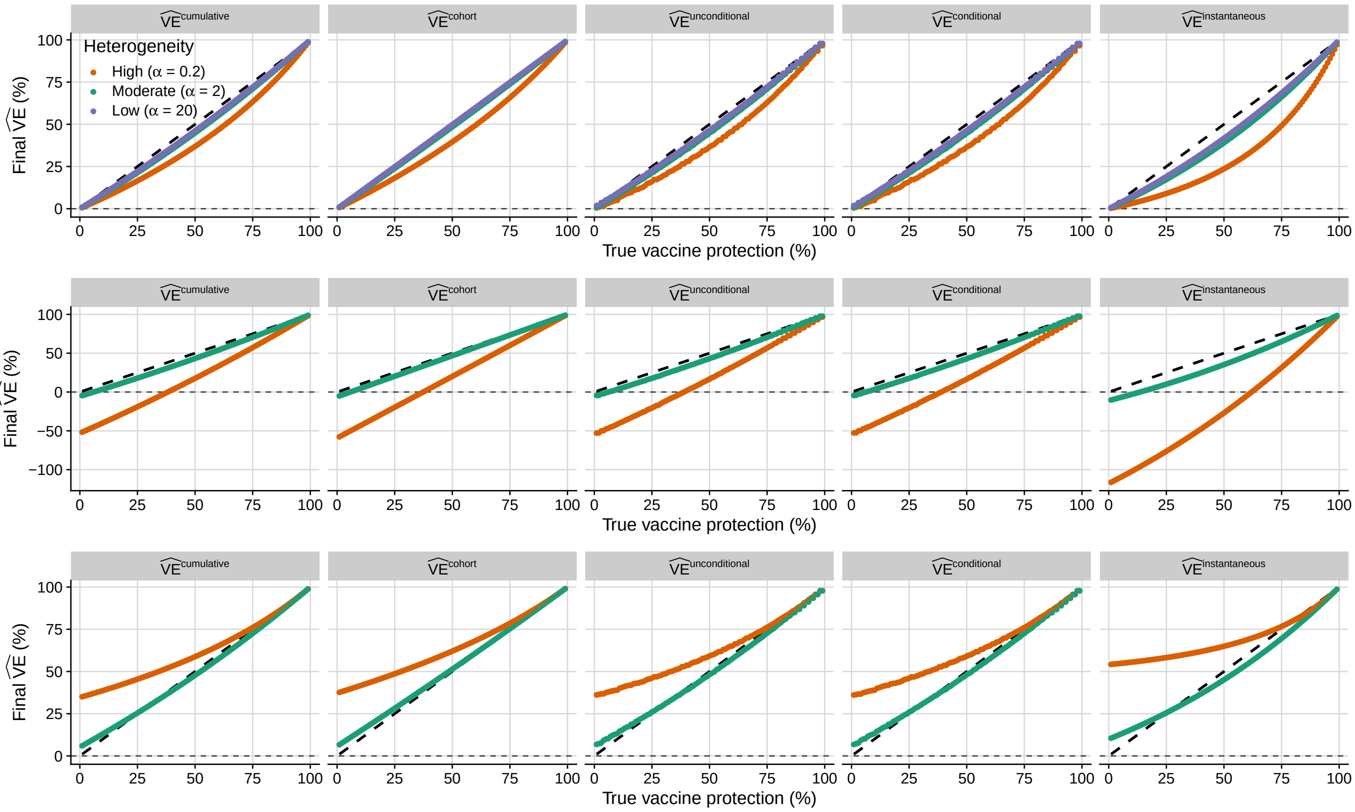

**Figure S.8:** Differential heterogeneity in pre-vaccination susceptibility affects final $\widehat{\mathrm{VE}}$ in the same direction, regardless how $\widehat{\mathrm{VE}}$ is estimated. Panel rows correspond to the same scenarios in Fig. 2: heterogeneity is identical in the vaccinated and unvaccinated populations (top row), heterogeneity is higher in the unvaccinated population relative to low heterogeneity ($\alpha_{\mathrm{v}} = 20$) in the vaccinated population (middle row), or heterogeneity is higher in the vaccinated population relative to low heterogeneity ($\alpha_{\mathrm{u}} = 20$) in the unvaccinated population (bottom row). The left-most panels, $\widehat{\mathrm{VE}}^{\mathrm{conditional}}$ estimates, are nearly identical to Fig. 2C, F, and I. Small differences between this figure and Fig. 2 are due to the shorter epidemic time to ensure an integral number of two-week intervals for regression-based $\widehat{\mathrm{VE}}$. Cohort and regression-based $\widehat{\mathrm{VE}}$ estimates vary in magnitude and direction similarly to $\mathrm{VE}^{\mathrm{cumulative}}$, though $\widehat{\mathrm{VE}}^{\mathrm{cohort}}$ estimates are closer to true protection when heterogeneity is the same in vaccinated and unvaccinated populations (top row). $\widehat{\mathrm{VE}}$ estimated from instantaneous incidence rates, $\widehat{\mathrm{VE}}^{\mathrm{instantaneous}}$ (right-most column), varies in the same direction but diverges more. All epidemics lasted for 196 days and parameter values are listed in Supplemental Table S.3.

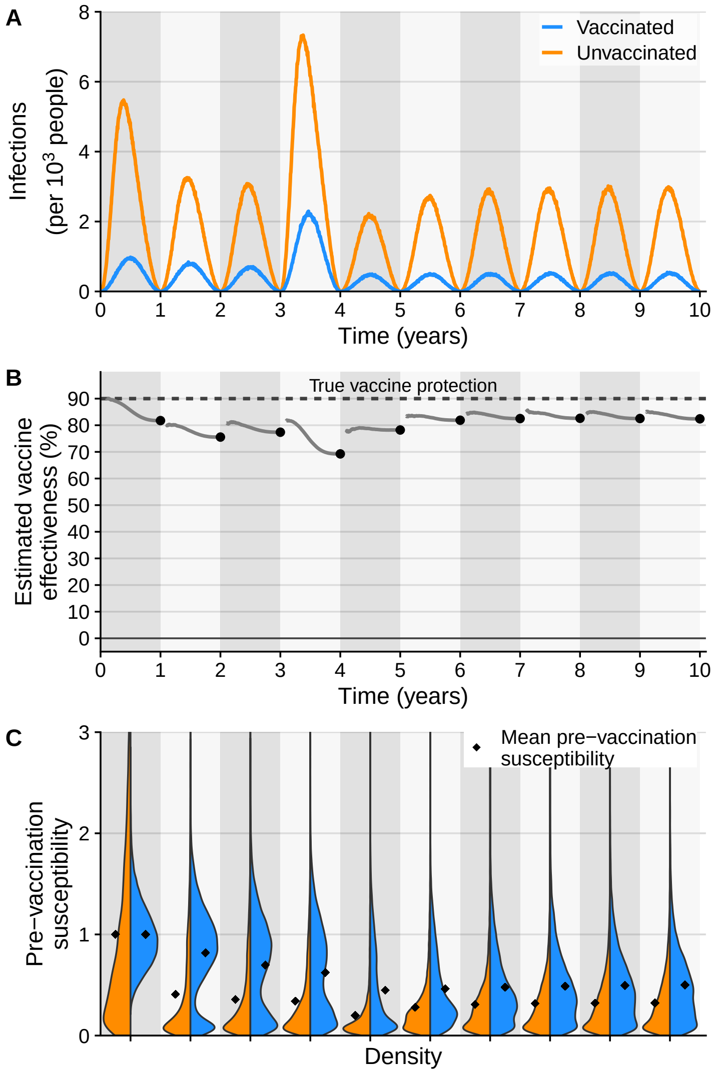

**Figure S.9:** Multiyear simulation with 90% true vaccine protection. All other simulation parameters are identical to the simulation used in Fig. 3.

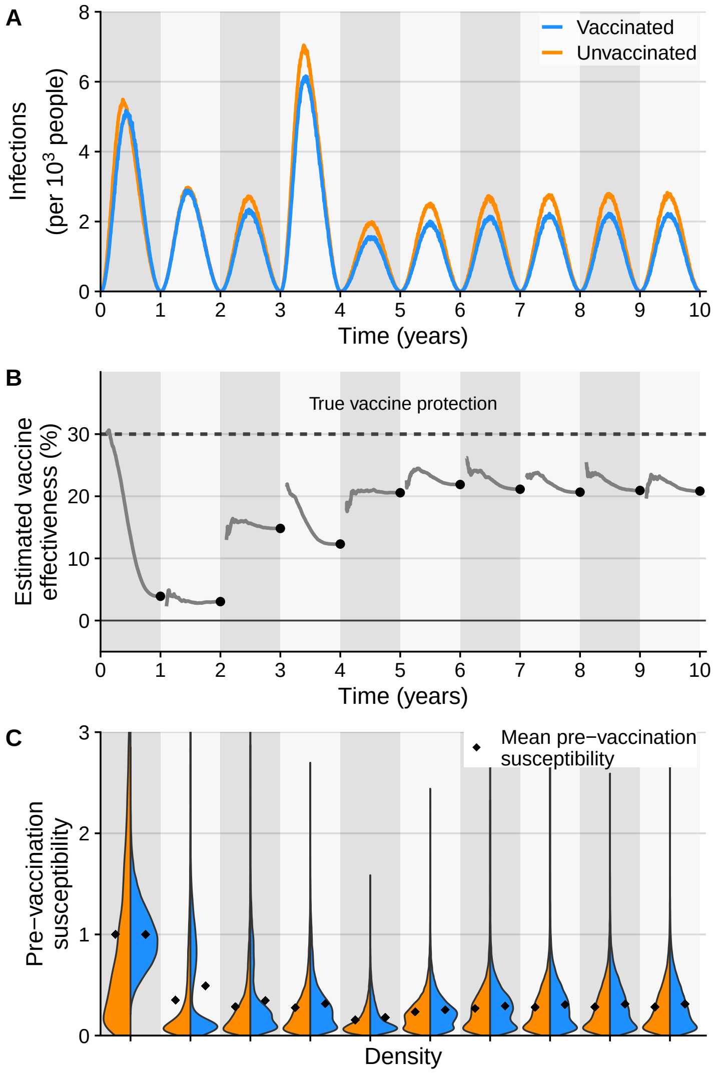

**Figure S.10:** Multiyear simulation with 30% true vaccine protection. All other simulation parameters are identical to the simulation used in Fig. 3.

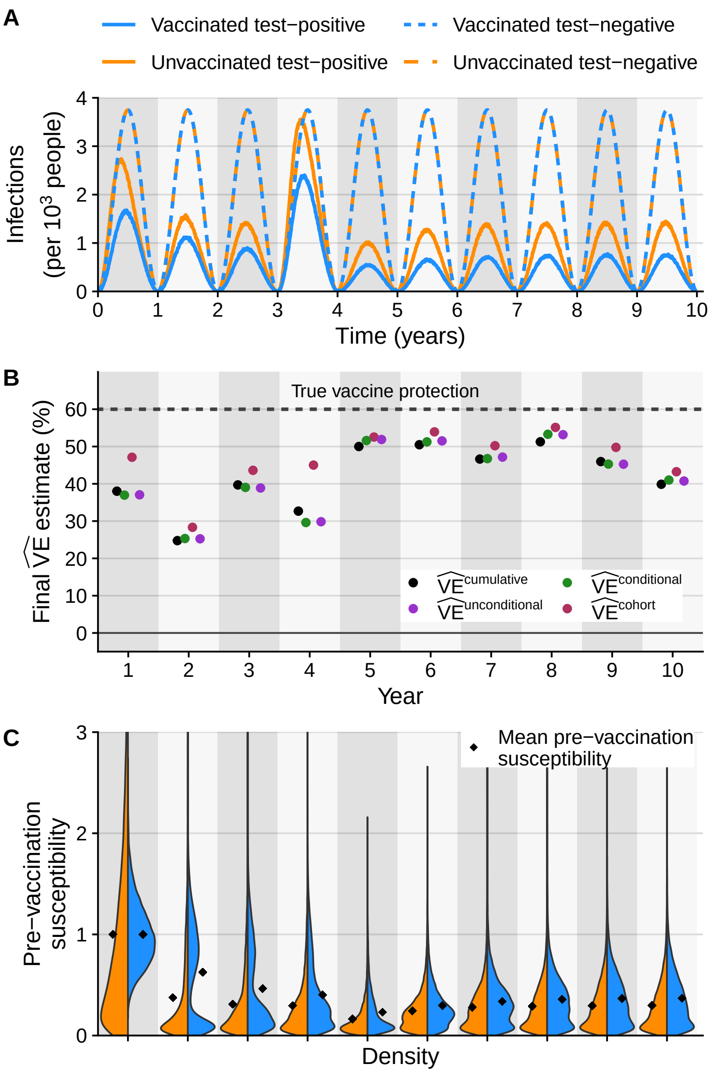

**Figure S.11:** Dynamic population immunity affects final annual $\widehat{\text{VE}}$ estimates regardless of how they are estimated. The test-positive infection incidence (solid curves in A) and pre-vaccination susceptibility distributions (C) are identical to Fig. 3. We estimated test-negative infections using the same seasonality as test-positive infections but with a higher cumulative attack rate (dashed curves in A). Final regression-based $\widehat{\text{VE}}$ estimates (purple and green dots in B) and $\widehat{\text{VE}}^{\text{cohort}}$ estimates (red dots in B) are shown alongside $\widehat{\text{VE}}^{\text{cumulative}}$ estimates (black dots in B) for each year. These final $\widehat{\text{VE}}^{\text{cumulative}}$ estimates differ slightly from those in Fig. 3 because a random sample of annual simulated infections is used for $\widehat{\text{VE}}$ calculations to ensure conditional regression models converge.

### References

[1] Joseph A Lewnard et al. "Measurement of Vaccine Direct Effects Under the Test-Negative Design". In: *American Journal of Epidemiology* 187.12 (Dec. 1, 2018), pp. 2686–2697. DOI: 10.1093/aje/kwy163.

[2] Ivo M. Foppa et al. "The case test-negative design for studies of the effectiveness of influenza vaccine". In: *Vaccine* 31.30 (June 26, 2013), pp. 3104–3109. DOI: 10.1016/j.vaccine.2013.04.026.

[3] P G Smith, L C Rodrigues, and P E M Fine. "Assessment of the Protective Efficacy of Vaccines against Common Diseases Using Case-Control and Cohort Studies". In: *International Journal of Epidemiology* 13.1 (1984), pp. 87–93. DOI: 10.1093/ije/13.1.87.

[4] Sylvia Ranjeva et al. "Age-specific differences in the dynamics of protective immunity to influenza". In: *Nature Communications* 10.1 (Apr. 10, 2019), p. 1660. DOI: 10.1038/s41467-019-09652-6.

[5] May P. S. Yeung, Frank L.Y. Lam, and Richard Coker. "Factors associated with the uptake of seasonal influenza vaccination in adults: a systematic review". In: *Journal of Public Health* 38.4 (Dec. 2, 2016), pp. 746–753. DOI: 10.1093/pubmed/fdv194.

[6] Philip Arevalo et al. "Earliest infections predict the age distribution of seasonal influenza A cases". In: *eLife* 9 (July 7, 2020). Ed. by Ben S Cooper et al., e50060. DOI: 10.7554/eLife.50060.

[7] Sean L. Wu et al. *Principled simulation of agent-based models in epidemiology*. Pages: 2020.12.21.423765 Section: New Results. Dec. 21, 2020. DOI: 10.1101/2020.12.21.423765.

[8] M. Lipsitch et al. "Depletion-of-susceptibles bias in influenza vaccine waning studies: how to ensure robust results". In: *Epidemiology & Infection* 147 (Jan. 2019), e306. DOI: 10.1017/S0950268819001961.
